# Assessing the Utility of ChatGPT Throughout the Entire Clinical Workflow

**DOI:** 10.1101/2023.02.21.23285886

**Authors:** Arya Rao, Michael Pang, John Kim, Meghana Kamineni, Winston Lie, Anoop K. Prasad, Adam Landman, Keith J Dreyer, Marc D. Succi

**Author notes:** **Corresponding Author:** Marc D. Succi, MD, Massachusetts General Hospital, Department of Radiology, 55 Fruit Street, Boston, MA, 02114, @MarcSucciMD. These authors contributed equally to this manuscript (Arya Rao, Michael Pang). **Data Sharing Statement:** All data generated or analyzed during the study are included in the published paper.

## Abstract

**IMPORTANCE:** Large language model (LLM) artificial intelligence (AI) chatbots direct the power of large training datasets towards successive, related tasks, as opposed to single-ask tasks, for which AI already achieves impressive performance. The capacity of LLMs to assist in the full scope of iterative clinical reasoning via successive prompting, in effect acting as virtual physicians, has not yet been evaluated.

**OBJECTIVE:** To evaluate ChatGPT’s capacity for ongoing clinical decision support via its performance on standardized clinical vignettes.

**DESIGN:** We inputted all 36 published clinical vignettes from the Merck Sharpe & Dohme (MSD) Clinical Manual into ChatGPT and compared accuracy on differential diagnoses, diagnostic testing, final diagnosis, and management based on patient age, gender, and case acuity.

**SETTING:** ChatGPT, a publicly available LLM

**PARTICIPANTS:** Clinical vignettes featured hypothetical patients with a variety of age and gender identities, and a range of Emergency Severity Indices (ESIs) based on initial clinical presentation.

**EXPOSURES:** MSD Clinical Manual vignettes

**MAIN OUTCOMES AND MEASURES:** We measured the proportion of correct responses to the questions posed within the clinical vignettes tested.

**RESULTS:** ChatGPT achieved 71.7% (95% CI, 69.3% to 74.1%) accuracy overall across all 36 clinical vignettes. The LLM demonstrated the highest performance in making a final diagnosis with an accuracy of 76.9% (95% CI, 67.8% to 86.1%), and the lowest performance in generating an initial differential diagnosis with an accuracy of 60.3% (95% CI, 54.2% to 66.6%). Compared to answering questions about general medical knowledge, ChatGPT demonstrated inferior performance on differential diagnosis (β=-15.8%, p<0.001) and clinical management (β=-7.4%, p=0.02) type questions.

**CONCLUSIONS AND RELEVANCE:** ChatGPT achieves impressive accuracy in clinical decision making, with particular strengths emerging as it has more clinical information at its disposal.

## Introduction

Despite its relative infancy, artificial intelligence (AI) is transforming healthcare, with current uses including workflow triage, predictive models of utilization, labeling and interpretation of radiographic images, patient support via interactive chatbots, communication aids for non-English speaking patients, and more.^1–8^ Yet, all of these use cases are limited to a specific part of the clinical workflow and do not provide longitudinal patient or clinician support. An under-explored use of AI in medicine is predicting and synthesizing patient diagnoses, treatment plans, and outcomes. Until recently, AI models have lacked sufficient accuracy and power to engage meaningfully in the clinical decision-making space. However, the advent of large language models (LLMs), which are trained on large amounts of human-generated text like the Internet, has motivated further investigation into whether AI can serve as an adjunct in clinical decision making throughout the entire clinical workflow, from triage to diagnosis to management. In this study, we assess the performance of a novel LLM, ChatGPT, on comprehensive clinical vignettes (short, hypothetical patient cases used to test clinical knowledge and reasoning).

ChatGPT is a popular chatbot derivative of OpenAI’s Generative Pre-trained Transformer-3.5 (GPT-3.5), an autoregressive large language model (LLM) released in 2022.^9^ Due to the chatbot’s widespread availability, a small but growing volume of preliminary studies have described ChatGPT’s performance on various professional exams (e.g. medicine, law, business, accounting)^10–14^ and generating highly technical texts as found in biomedical literature.^15^ Recently, there has been great interest in utilizing the nascent but powerful chatbot for clinical decision support.^16–18^

Given that LLMs like ChatGPT have the ability to integrate large amounts of textual information to synthesize responses to human-generated prompts, we speculated that ChatGPT would be able to act as an on-the-ground copilot in clinical reasoning, making use of the wealth of information available during patient care from the Electronic Health Record (EHR) and other sources. We focused on comprehensive clinical vignettes as a model, and tested the hypothesis that when provided clinical vignettes, ChatGPT would be able to recommend diagnostic workup, decide the clinical management course, and ultimately make the diagnosis, working through the entire clinical encounter.

Our study is the first to make use of ChatGPT’s ability to integrate information from the earlier portions of a conversation into downstream responses. Thus, this model lends itself well to the iterative nature of clinical medicine, in that the influx of new information requires constant updating of prior hypotheses.

## Methods

### Study Design

We assessed ChatGPT’s accuracy in solving comprehensive clinical vignettes, comparing across patient age, gender, and acuity of clinical presentation. We presented each portion of the clinical workflow as a successive prompt to the model (differential diagnosis, diagnostic testing, final diagnosis, and clinical management questions were presented one after the other) (Figure 1A).

**Figure 1:**
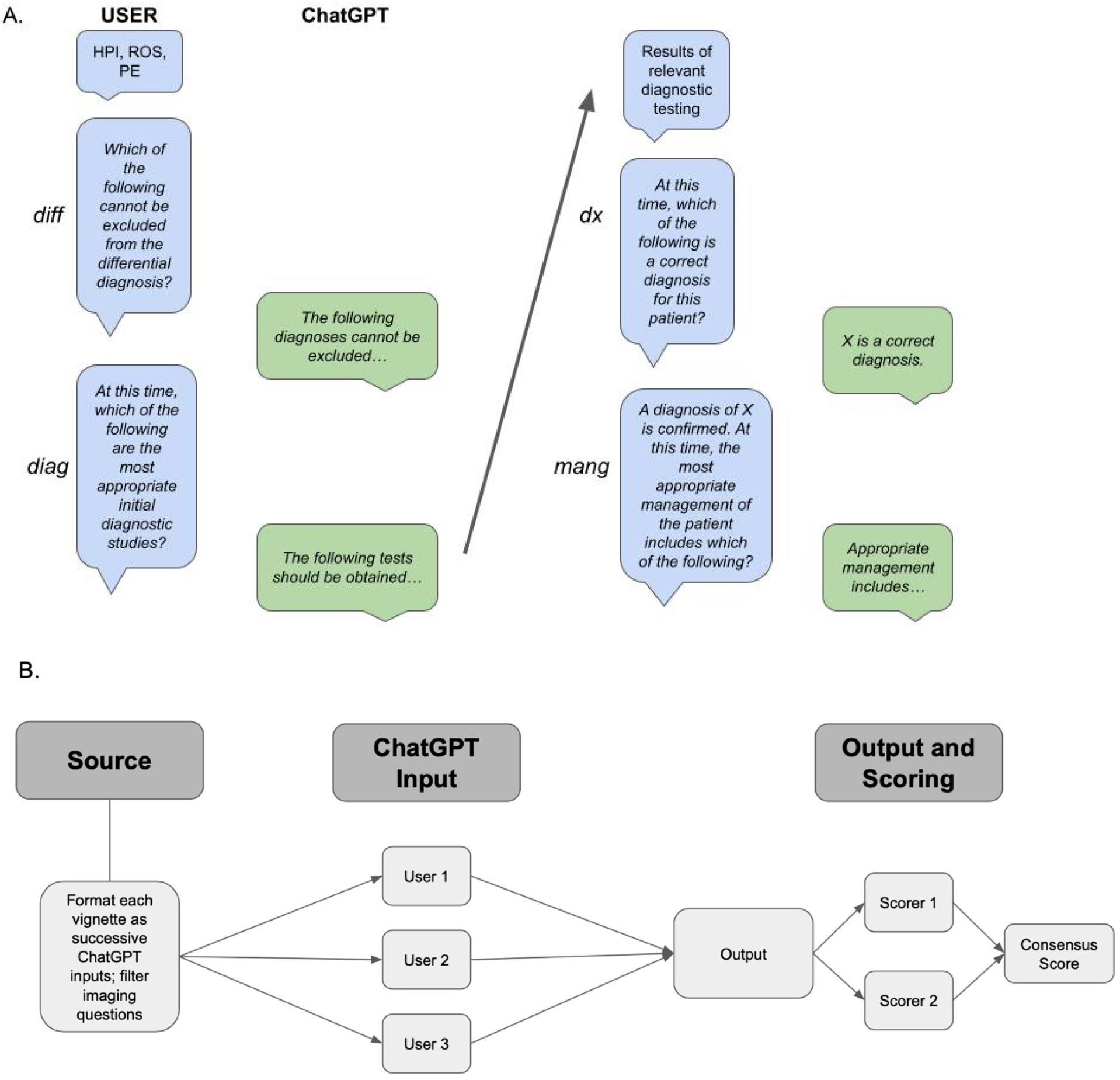
Experimental workflow for determining ChatGPT accuracy in solving clinical vignettes. **Panel A**: Schematic of user interface with ChatGPT for this experiment. Blue boxes indicate prompts given to ChatGPT and green boxes indicate ChatGPT responses. Non-italicized text indicates information given to ChatGPT without a specific question attached. **Panel B:** Schematic of experimental workflow. Prompts were developed from MSD vignettes and converted to ChatGPT-compatible text input. Questions requiring the interpretation of images were removed. Three independent users tested each prompt. Two independent scorers calculated scores for all outputs; these were compared to generate a consensus score.

### Setting

ChatGPT (San Francisco, OpenAI) is a transformer-based language model with the ability to generate human-like text. It captures the context and relationship between words in input sequences through multiple layers of self-attention and feed-forward neural networks. The language model is trained on a variety of text including websites, articles, and books up until 2021. The ChatGPT model is self-contained in that it does not have the ability to search the internet when generating responses. Instead, it predicts the most likely “token” to succeed the previous one based on patterns in its training data. Therefore, it does not explicitly search through existing information, nor does it copy existing information. All ChatGPT model output was collected from the January 9, 2023 version of ChatGPT.

### Data Sources and Measurement

Clinical vignettes were selected from the Merck Sharpe & Dohme Clinical Manuals, referred to as the MSD Manuals^19^. These vignettes represent canonical cases that commonly present in healthcare settings and include components analogous to clinical encounter documentation such as the history of present illness (HPI), review of systems (ROS), physical exam (PE), and laboratory test results. The vignette online modules include sequential “select all that apply” (SATA) type questions to simulate differential diagnosis, diagnostic workup, and clinical management decisions. They are written by independent experts in the field and undergo a peer review process before being published. At the time of the study, 36 vignette modules were available online, and 34 of the 36 were available online as of ChatGPT’s September 2021 training cutoff. All 36 modules passed the eligibility criteria and were included in the ChatGPT model assessment.

Case transcripts were generated by copying MSD manual vignettes directly into ChatGPT. Questions posed in the MSD manual vignettes were presented as successive inputs to ChatGPT (Figure 1B). All questions requesting the clinician to analyze images were excluded from our study, as ChatGPT is a text-based AI without the ability to interpret visual information.

ChatGPT’s answers are informed by the context of the ongoing conversation. To avoid the influence of other vignettes’ answers on model output, a new ChatGPT session was instantiated for each vignette. A single session was maintained for each vignette and for all associated questions, allowing ChatGPT to take all available vignette information into account as it proceeds to answer new questions. To account for response-by-response variation, each vignette was tested in triplicate, each time by a different user. Prompts were not modified from user to user.

We awarded points for each correct answer given by ChatGPT and noted the total number of correct decisions possible for each question. For example, for a question asking whether each of a list of diagnostic tests is appropriate for the patient presented, a point was awarded for each time ChatGPT’s answer was concordant with the provided Merck answer.

Two scorers independently calculated an individual score for each output to ensure consensus on all output scores; there were no scoring discrepancies. The final score for each prompt was calculated as an average of the three replicate scores. Based on the total possible number of correct decisions per question, we calculated a proportion of correct decisions for each question (“average proportion correct” refers to the average proportion across replicates). A schematic of the workflow is provided in Figure 1A.

### Participants and Variables

The MSD vignettes feature hypothetical patients and include information on the age and gender of each patient. We used this information to assess the effect of age and gender on accuracy. To assess differential performance across the range of clinical acuity, the Emergency Severity Index (ESI)^20^ was used to rate the acuity of the MDS clinical vignettes. The ESI is a five-level triage algorithm to assign patient priority in the emergency department. Assessment is based on medical urgency and assesses the patient’s chief complaint, vital signs, and ability to ambulate. The ESI is an ordinal scale ranging from 1 to 5 corresponding to highest to lowest acuity respectively. For each vignette, we fed the HPI into ChatGPT to determine its ESI and cross-validated with human ESI scoring. All vignette metadata, including title, age, gender, ESI, and final diagnosis, can be found in eTable1.

Questions posed by the MSD Manual vignettes fall into several categories: differential diagnoses (abbreviated as *diff*) which ask the user to determine which of several conditions cannot be eliminated from an initial differential, diagnostic questions (abbreviated as *diag*) which ask the user to determine appropriate diagnostic steps based on the current hypotheses and information, diagnosis questions (abbreviated as *dx*) which ask the user for a final diagnosis, management questions (abbreviated as *mang*) which ask the user to recommend appropriate clinical interventions, and miscellaneous questions (abbreviated as *misc*) which ask the user medical knowledge questions relevant to the vignette, but not necessarily specific to the patient at hand. We stratified results by question type and the demographic information previously described.

### Statistical Methods

Multivariable linear regression was performed using the lm() function with R version 4.2.1 (Vienna, R Core Team) to assess the relationship between ChatGPT vignette performance, question type, demographic variables (age, gender), and clinical acuity (ESI). Question type was dummy-variable-encoded to assess the effect of each category independently. The *misc* question type was chosen as the reference variable as these questions assess general knowledge and not necessarily active clinical reasoning. Age, gender, and ESI were also included in the model to control for potential sources of confounding.

## Results

### Overall Performance

Since questions from all vignettes fall into several distinct categories, we were able to assess performance not only on a vignette-by-vignette basis, but also on a category-by-category basis. We found that on average, across all vignettes, ChatGPT achieved 71.8% accuracy (Figure 2A, eTable 2, eTable3). Between categories and across all vignettes, ChatGPT achieved the highest accuracy (76.9%) for questions in the *dx* category, and the lowest accuracy for questions in the *diff* category (60.3%) (Figure 2B, eTable3). Trends for between-question-type variation in accuracy for each vignette are shown in Figure 2C.

**Figure 2:**
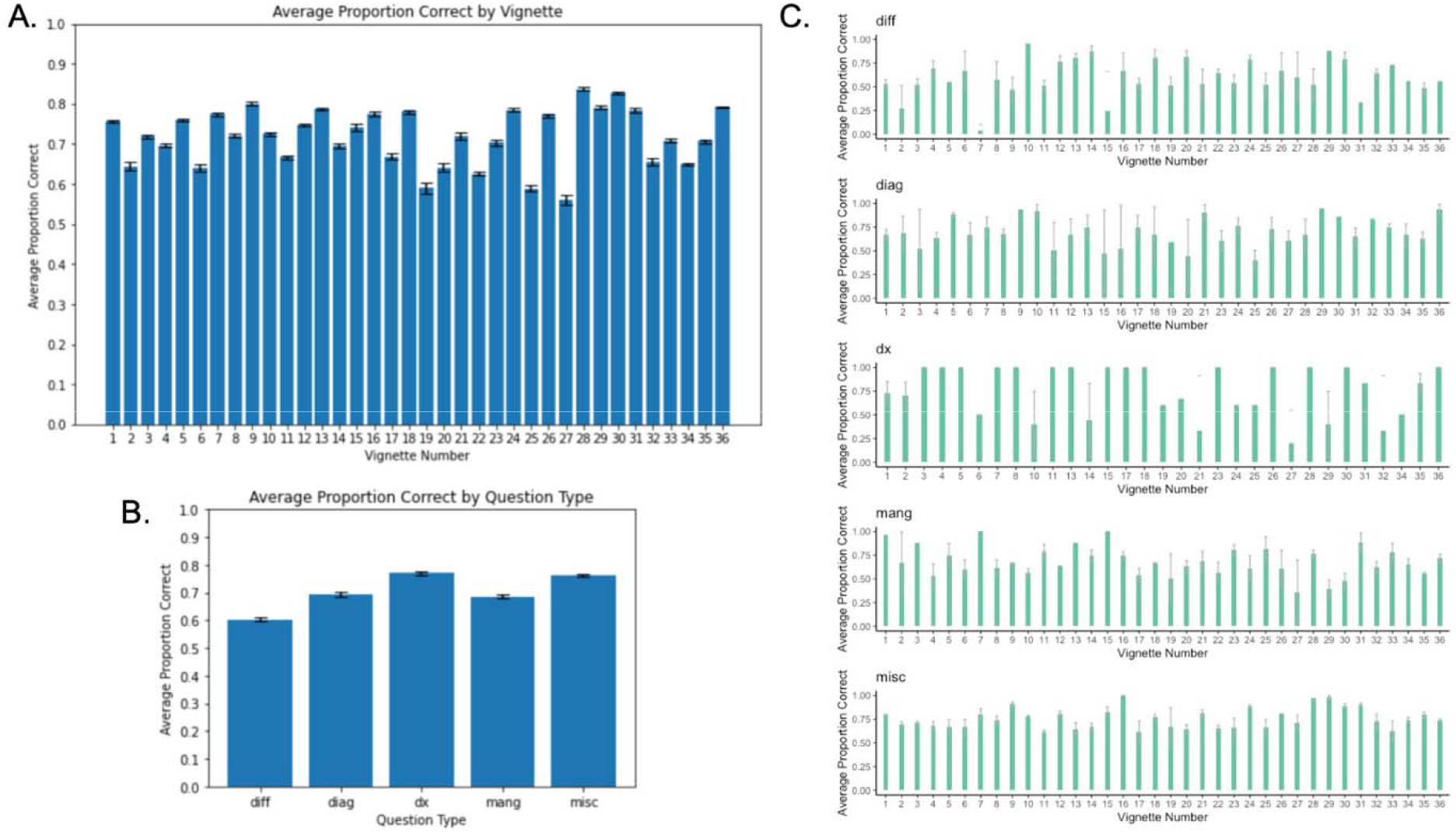
ChatGPT performance on clinical vignettes by vignette and by question type. **Panel A:** ChatGPT overall performance for each of the 36 MSD vignettes; error bars are +/- 1 standard error of the mean. **Panel B:** ChatGPT performance by question type; error bars are +/- 1 standard error of the mean. **Panel C:** ChatGPT performance by question type for each of the 36 MSD vignettes; error bars are +/- 1 standard error of the mean.

Vignette #28, featuring a right testicular mass in a 28-year-old man (final diagnosis of testicular cancer), showed the highest accuracy overall (83.8%). Vignette #27, featuring recurrent headaches in a 31-year-old woman (final diagnosis of pheochromocytoma), showed the lowest accuracy overall (55.9%) (Figure 2A, eTable2).

### Differential Versus Final Diagnosis

*Diff* and *dx* questions ask the user to generate a broad differential diagnosis followed by a final diagnosis respectively. The key difference between the two question types is that answers to *diff* questions rely solely on the HPI, ROS, and PE, while answers to *dx* questions incorporate results from relevant diagnostic testing and potentially additional clinical context. Therefore, a comparison between the two sheds light on whether ChatGPT’s utility in the clinical setting improves with the amount of accurate, patient-specific information it has access to.

We found a statistically significant difference in performance between these two question types overall (Figure 2B). Average performance on *diff* questions was 60.3%, and average performance on *dx* questions was 76.9%, indicating a 16.6% average increase in accuracy in diagnosis as more clinical context is provided. We also found that there were statistically significant differences in accuracy between *diff* and *dx* questions within vignettes for the majority of vignettes. This indicates that this is not an aggregate phenomenon, but rather one that applies broadly (Figure 2C).

### Performance Across Patient Age and Gender

The MSD vignettes specify both the age and gender of patients. We performed multivariable linear regression analysis to investigate the effect of age and gender on ChatGPT accuracy. Regression coefficients for age and gender were both not significant (Table 1). This result suggests ChatGPT performance is equivalent across the range of ages in this study as well as in a binary definition of gender.

**Table 1:**
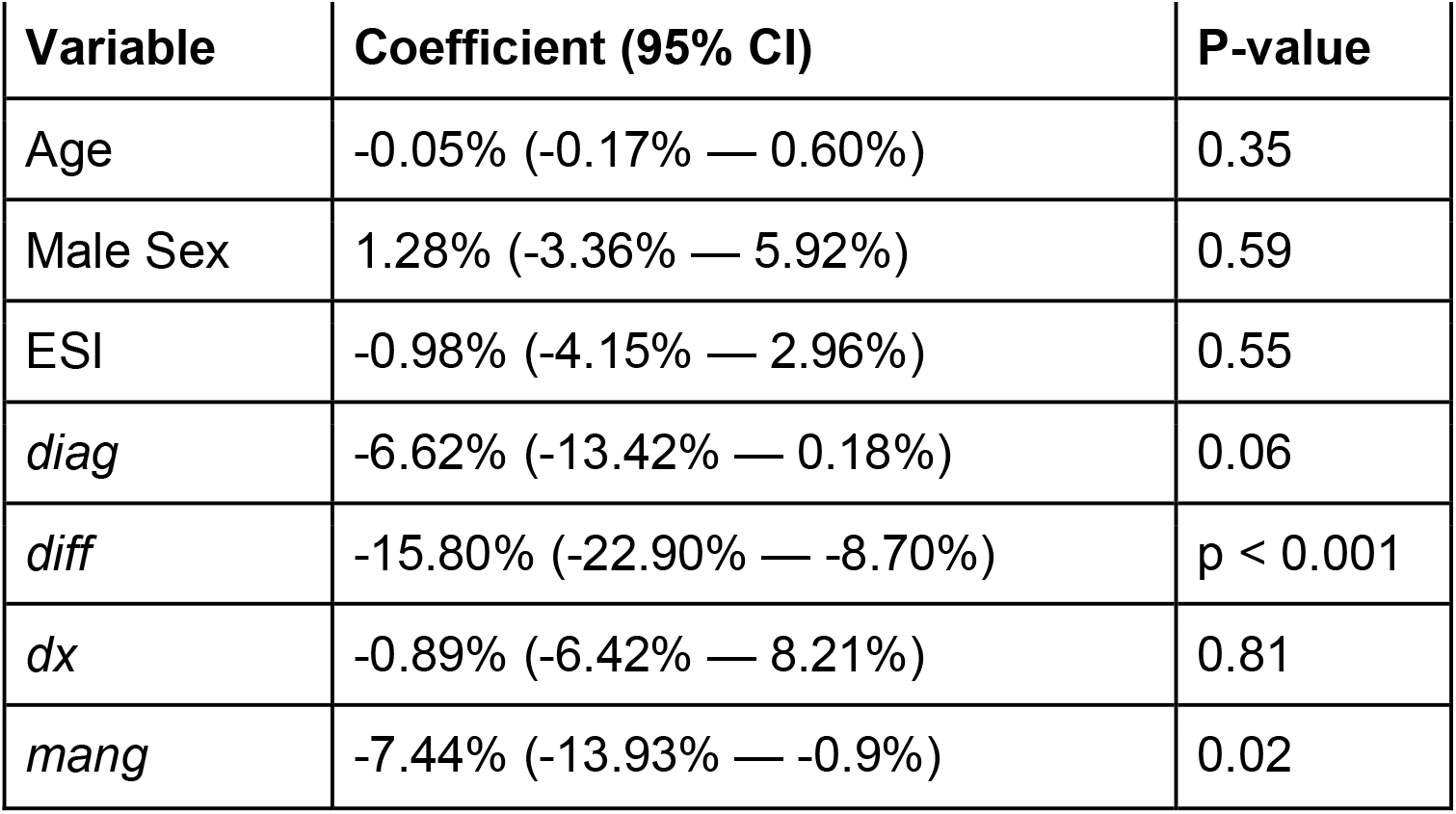
Results of multivariable linear regression examining the relationship between ChatGPT accuracy and the age, sex, ESI of the clinical vignette patient as well as the question type.

| Variable | Coefficient (95% CI) | P-value |
| --- | --- | --- |
| Age | -0.05% (-0.17% — 0.60%) | 0.35 |
| Male Sex | 1.28% (-3.36% — 5.92%) | 0.59 |
| ESI | -0.98% (-4.15% — 2.96%) | 0.55 |
| *diag* | -6.62% (-13.42% — 0.18%) | 0.06 |
| *diff* | -15.80% (-22.90% — -8.70%) | p < 0.001 |
| *dx* | -0.89% (-6.42% — 8.21%) | 0.81 |
| *mang* | -7.44% (-13.93% — -0.9%) | 0.02 |

### ChatGPT Performance Across Question Types

Differential and management type questions were negatively associated with ChatGPT performance relative to miscellaneous type questions (β=-15.8%, p<0.001 and β=-7.4%, p=0.02 respectively). Diagnostic questions trended towards decreased performance, however, the effect was not statistically significant. There was no difference in performance in final diagnosis accuracy. The R-squared value of the model was 0.083 indicating that only 8.3% of the variance in ChatGPT accuracy was explained by the model. This suggests that other factors may play a role in explaining variation in ChatGPT performance.

### ChatGPT Performance Does Not Vary With Acuity of Clinical Presentation

Case acuity was assessed by asking ChatGPT to provide the ESI for each vignette based on only the HPI. These ratings were validated for accuracy by human scorers. ESI was included as an independent variable in the multivariable linear regression shown in Table 1, but was not a significant predictor of ChatGPT accuracy.

### ChatGPT Performance is Ambiguous with Respect to Dosing of Medications

A small subset of *mang* and *misc* questions demanded that ChatGPT provide numerical answers such as dosing for particular medications. Qualitative analysis of ChatGPT’s responses indicates that errors in this subset are predisposed towards incorrect dosing rather than incorrect medication (eTable 4).

## Discussion

In this study, we present first-of-its-kind evidence assessing the potential use of novel artificial intelligence tools throughout the entire clinical workflow, encompassing initial diagnostic workup, diagnosis, and clinical management. We provide the first analysis of ChatGPT’s iterative prompt functionality in the clinical setting, reflecting the constantly shifting nature of patient care by allowing upstream prompts and responses to affect downstream answers. We show that ChatGPT achieves 60.3% accuracy in determining differential diagnoses based on the HPI, PE, and ROS alone. With additional information such as the results of relevant diagnostic testing, ChatGPT achieves 76.9% accuracy in narrowing a final diagnosis.

ChatGPT achieves an average performance of 71.8% across all vignettes and question types. Notably, of the patient-focused questions posed by each vignette, ChatGPT achieved the highest accuracy (76.9% on average) answering *dx* questions, which prompted the model to provide a final diagnosis based on HPI, PE, ROS, diagnostic results, and any other pertinent clinical information. There was no statistical difference between *dx* accuracy and *misc* accuracy, indicating that ChatGPT performance on a specific clinical case, when provided with all possible relevant clinical information, approximates its accuracy in providing general medical facts.

Overall accuracy was lower for *diag* and *mang* questions than for *diff* and *dx* questions (Figure 2B). In some cases, this was because ChatGPT recommended extra or unnecessary diagnostic testing or clinical intervention, respectively (eTable 4). In contrast, for several *diff* and *dx* questions (for which all necessary information was provided to answer, as for the *diag* and *mang* questions), ChatGPT refused to provide a diagnosis altogether (eTable 4). This indicates ChatGPT is not always able to properly navigate clinical scenarios with a well-established standard of care (ex. a clear diagnosis based on a canonical presentation) and situations in which the course of action is more ambiguous (ex. ruling out unnecessary testing). The latter observation is in line with Rao et al.’s observation that ChatGPT struggles to identify situations in which diagnostic testing is futile.^17^ Resource utilization was not explicitly tested in our study; further prompt engineering could be performed to evaluate ChatGPT’s ability to recommend the appropriate utilization of resources (for example, asking “What tests are appropriate clinically while also taking cost management into account?”).

Rao et al. found that for breast cancer and breast pain screening, ChatGPT’s accuracy in determining appropriate radiologic diagnostic workup varied with the severity of initial presentation. For breast cancer, there was a positive correlation between severity and accuracy, and for breast pain there was a negative correlation.^17^ Given that the data in this study covers 36 different clinical scenarios as opposed to trends within specific clinical conditions, we suspect that any association between acuity of presentation and accuracy could be found on a within-case basis, as opposed to between cases.

Given the important ongoing discourse^3–8^ surrounding bias in the clinical setting and bias in artificial intelligence, we believe our analysis of ChatGPT’s performance based on the age and gender of patients represents an important touchpoint in both discussions.^21–25^ While we did not find that age or gender is a significant predictor of accuracy, we note that our vignettes represent classic presentations of disease, and that atypical presentations may generate different biases. Further investigation into additional demographic variables and possible sources of systematic bias is warranted in future studies.

While on the surface ChatGPT performs impressively, it is worth noting that even small errors in clinical judgment can result in adverse outcomes. ChatGPT’s answers are generated based on finding the next most likely “token” or word/phrase to complete the ongoing answer; as such, ChatGPT lacks reasoning capacity. This is evidenced by instances in which ChatGPT recommends futile care or refuses to provide a diagnosis even when equipped with all necessary information and is further evidenced by its frequent errors in dosing. These limitations are inherent to the artificial intelligence model itself and can be broadly divided into several categories, including misalignment and hallucination.^26,27^ In this study, we identified and accounted for these limitations with replicate validation. These considerations are necessary when determining both the parameters of artificial intelligence utilization in the clinical workflow and the regulations surrounding the approval of similar technologies in clinical settings.

As applications of AI grow more ubiquitous in every sector, it is important to not only understand if such tools are reliable in the clinical setting, but also to postulate the most effective methods for deploying them. By analyzing ChatGPT’s accuracy not just at one step, but rather throughout the entire clinical workflow, our study provides a realistic pilot of how LLMs like ChatGPT might perform in the clinical settings. Integration of LLMs with existing EHR (with appropriate regulations) could facilitate improved patient outcomes and workflow efficiency.

## Data Availability

All data produced in the present work are contained in the manuscript

