## Supplementary Tables for "Assessing the Utility of ChatGPT Throughout the Entire Clinical Workflow"

**eTable1: Metadata for MSD Vignettes**

| **Vignette Number** | **Title** | **Age** | **Gender** | **ESI** | **Final Diagnosis** |
| --- | --- | --- | --- | --- | --- |
| 1 | Abdominal Pain in a 26-Yr-Old Pregnant Woman | 26 | F | 3 | Sepsis |
| 2 | Anemia in a 42-year-old woman | 42 | F | 2 | Transient pure red cell aplasia triggered by parvovirus B19 infection |
| 3 | Back Discomfort in a 12-Year-Old Boy | 12 | M | 4 | Adolescent idiopathic scoliosis |
| 4 | Chest Pain in 74-Yr-Old Man | 74 | M | 2 | Non-ST-segment elevation myocardial infarction (NSTEMI) |
| 5 | Chest Pain in a 49-Yr-Old Woman | 49 | F | 2 | Pulmonary embolism (PE) |
| 6 | Chest Pain in a 62-Yr-Old Man | 62 | M | 4 | Stable angina |
| 7 | Chronic Lethargy in a 68-Yr-Old Woman | 68 | F | 3 | Hashimoto thyroiditis |
| 8 | Confusion in a 74-Yr-Old Woman | 74 | F | 2 | Seizure due to meningioma |
| 9 | Cough in 47-Yr-Old Woman | 47 | F | 2 | Asthma |
| 10 | Cough in a 2-Yr-Old Boy | 2 | M | 3 | Foreign body aspiration, left lung |
| 11 | Cough in a 54-Yr-Old Man | 54 | M | 3 | Community-acquired pneumonia |
| 12 | Dyspnea and Back Pain in a 24-Yr-Old Man | 24 | M | 3 | Primary spontaneous pneumothorax |
| 13 | Exertional Dyspnea in a 76 Yr-Old Man | 76 | M | 2 | Severe aortic stenosis |
| 14 | Fatigue and Palpitations in a 53-Yr-Old Woman | 53 | F | 3 | Atrial fibrillation |
| 15 | Headache and dizziness in a 54-year-old man | 54 | M | 4 | Acoustic neuroma, right side |
| 16 | Hyperglycemia in a 56-Year-Old Woman | 56 | F | 4 | Type 2 diabetes |
| 17 | Hypernasalilty and Ptosis in a 48-Yr-Old Woman | 48 | F | 2 | Myasthenia gravis |
| 18 | Left Scrotal Pain in an 18-Yr-Old Man | 18 | M | 3 | Testicular torsion |
| 19 | Left-sided Abdominal Pain in a 45-Yr-Old Man | 45 | M | 3 | Ureteral lithiasis |
| 20 | Leg Pain in a 62-Yr-Old Man | 62 | M | 3 | Cellulitis |
| 21 | Leg Tingling in 20-Yr-Old Woman | 20 | F | 3 | Multiple sclerosis |
| 22 | Lethargy in a 65-Yr-Old Man | 65 | M | 1 | Beta-blocker overdose |
| 23 | Loss of Consciousness in a 50-Yr-Old Man | 50 | M | 2 | Immune thrombocytopenia (ITP) |
| 24 | Nausea and Increased Thirst in a 31-Yr-Old Woman | 31 | F | 2 | Primary hyperparathyroidism due to PTH-secreting parathyroid adenoma |
| 25 | Nipple discharge in a 31-Yr-Old Woman | 31 | F | 4 | Galactorrhea |
| 26 | Nocturia in a 70-Yr-Old Man | 70 | M | 2 | Benign prostatic hyperplasia (BPH) |
| 27 | Recurrent Headaches in a 31-Yr-Old Woman | 31 | F | 2 | Pheochromocytoma |
| 28 | Right Testicular Mass in a 28-Yr-Old Man | 28 | M | 3 | Testicular cancer |
| 29 | Scrotal pain in a 10-Yr-Old Boy | 10 | M | 2 | Torsion of the testicular appendage |
| 30 | Scrotal Pain in a 21-Yr-Old Man | 21 | M | 3 | Epididymitis or orchitis |
| 31 | Shortness of Breath in a 68-Yr-Old Man | 68 | M | 2 | Restrictive cardiomyopathy |
| 32 | Syncope in an 80-Yr-Old Woman | 80 | F | 2 | Cardiac tamponade |
| 33 | Vaginal bleeding in a pregnant 32-year-old woman | 32 | F | 2 | Placental abruption |
| 34 | Weight Gain and Muscle Weakness in a 54-Yr-Old Man | 54 | M | 3 | Pituitary ACTH secretion (Cushing's disease) |
| 35 | Weight loss and dizziness in a 29-year-old man | 29 | M | 2 | Primary adrenal insufficiency |
| 36 | Weight Loss and Palpitations in a 41-Yr-Old Man | 41 | M | 3 | Graves disease |

Metadata for MSD vignettes. Age, gender, title, and final diagnosis were all provided within the vignettes themselves. ESI rating was calculated by ChatGPT and human scorers (see Methods). Vignette number was assigned by this research group in the order that the vignettes are published online.

**eTable2: ChatGPT Accuracy by Vignette**

| **Vignette Number** | **Average Proportion Correct** | **Standard Error of the Mean** |
| --- | --- | --- |
| 1 | 0.75634921 | 0.00235725 |
| 2 | 0.64326599 | 0.01073817 |
| 3 | 0.71883267 | 0.00458918 |
| 4 | 0.6957294 | 0.00446449 |
| 5 | 0.75984848 | 0.00426538 |
| 6 | 0.64074074 | 0.00878604 |
| 7 | 0.77407407 | 0.00494539 |
| 8 | 0.72117304 | 0.00479085 |
| 9 | 0.80092593 | 0.00353047 |
| 10 | 0.72460317 | 0.00459257 |
| 11 | 0.66684704 | 0.00419683 |
| 12 | 0.74772727 | 0.003634 |
| 13 | 0.78703704 | 0.00358782 |
| 14 | 0.69478438 | 0.00701748 |
| 15 | 0.74177489 | 0.00964328 |
| 16 | 0.77492151 | 0.00722724 |
| 17 | 0.66887755 | 0.00703032 |
| 18 | 0.77993197 | 0.00442806 |
| 19 | 0.58979107 | 0.01423041 |
| 20 | 0.64087302 | 0.01071508 |
| 21 | 0.7209127 | 0.00871775 |
| 22 | 0.6261992 | 0.00503332 |
| 23 | 0.70305217 | 0.00780044 |
| 24 | 0.78562031 | 0.00347551 |
| 25 | 0.58941799 | 0.00740824 |
| 26 | 0.77089947 | 0.00493125 |
| 27 | 0.55929705 | 0.0123026 |
| 28 | 0.83767952 | 0.00440924 |
| 29 | 0.79166667 | 0.00552734 |
| 30 | 0.82738095 | 0.00437194 |
| 31 | 0.78439153 | 0.00555443 |
| 32 | 0.65509259 | 0.00879226 |
| 33 | 0.70833333 | 0.00491073 |
| 34 | 0.64839958 | 0.00351188 |
| 35 | 0.70652054 | 0.00408724 |
| 36 | 0.79298777 | 0.00163206 |

Average proportion correct for each MSD vignette and associated standard error of the mean.

**eTable3: ChatGPT Accuracy by Question Type**

| **Question Type** | **Average Proportion Correct** | **Standard Error of the Mean** |
| --- | --- | --- |
| *diff* | 0.60354963 | 0.00577072 |
| *diag* | 0.69336524 | 0.00969417 |
| *dx* | 0.76975309 | 0.00697913 |
| *mang* | 0.68579747 | 0.00548833 |
| *misc* | 0.76129811 | 0.00465247 |

Average proportion correct for each question type across all MSD vignettes and associated standard error of the mean.

**eTable4: ChatGPT Raw Output**

| **Question ID** | **Output 1** | **Output 2** | **Output 3** |
| --- | --- | --- | --- |
| M1_diff | Based on the information provided, the differential diagnoses that cannot be excluded are:  Ectopic pregnancy Placental abruption Pelvic inflammatory disease (PID) Sepsis Septic abortion It is important to note that this is not a comprehensive list and further investigation and tests, such as imaging studies and laboratory tests, may be needed to confirm a diagnosis. The patient should receive prompt medical attention and evaluation. | At this time, the following differential diagnoses cannot be excluded:  Ectopic pregnancy Pelvic inflammatory disease (PID) Placental abruption Pyelonephritis Septic abortion Note: Further evaluation, such as laboratory tests, imaging studies and possibly an obstetrical consultation, is needed to determine the cause of the patient's symptoms. | At this time, the differential diagnoses cannot be excluded are:  Ectopic pregnancy Placental abruption Pelvic inflammatory disease (PID) Pyelonephritis Septic abortion Sepsis |
| M1_diag | Based on the information provided, the most appropriate initial diagnostic studies are:  Complete blood count (CBC) Blood culture Urinalysis and culture Serum beta-hCG Ultrasonography of the abdomen These tests can help to evaluate the patient's overall health, rule out certain infections and confirm pregnancy, and evaluate the uterus and adnexa for any abnormalities. However, this is not a comprehensive list and other tests may be needed based on the patient's individual case and any changes in her condition. The patient should receive prompt medical attention and evaluation. | Ultrasonography of the abdomen and pelvis, serum beta-hCG, blood culture, urinalysis and culture. | The most appropriate initial diagnostic studies are:  Complete blood count (CBC) Blood culture Urinalysis and culture Serum beta-hCG Ultrasonography of the kidney and bladder |
| M1_dx | Based on the information provided, a correct diagnosis for this patient would be:  Acute Pyelonephritis Cystitis | Acute Pyelonephritis | Acute pyelonephritis |
| M1_mang | Admit to the medical floor, IV Ceftriaxone 1 gm once a day, and IV 0.9% saline 500 mL bolus. | Admit to the medical floor, Ceftriaxone 1 gm IV once a day, IV 0.9% saline 500 mL bolus, IV dextrose 5% ¼ normal saline, 75 mL/hour. | Admit to medical floor, Ceftriaxone 1 gm IV once a day, IV 0.9% saline 500 mL bolus. |
| M1_q1 | E. coli is the most likely cause of pyelonephritis in this patient. | E. coli is the most likely cause of pyelonephritis in this patient. | E. coli is the most likely cause of pyelonephritis in this patient. |
| M1_q2 | All pregnant patients. | All pregnant patients. | All pregnant patients |
| M1_q3 | Temperature > 38° C, heart rate > 110 beats/minute, respiratory rate > 18/minute, and presence of infection are included in the criteria for sepsis. | Temperature > 38° C, Heart rate > 110 beats/minute, Respiratory rate > 18/minute, Presence of infection | Temperature > 38° C, Heart rate > 110 beats/minute, Respiratory rate > 18/minute, Presence of infection |
| M2_DIFF | The patient is a 42-year-old female with significant anemia, dyspnea on exertion, and fatigue. She was diagnosed with anemia a week prior at an urgent care center and has a CBC result of Hemoglobin=7.2 g/dL, RBC=3.5 million/cm3, WBC=7500/mm3, RDW=13, MCV=89 fL. She had the flu 2 months ago with diffuse joint pain and a non-pruritic rash over her extremities. She is a vegetarian, has no history of COPD, asthma, or lung disease, and has a history of mild anemia. She is a school teacher, swims regularly but has not done so lately, and is married and trying to conceive. She has had a COVID-19 vaccine. Physical exam shows no visible signs of distress, normal vital signs, clear lungs, and a slightly enlarged spleen. | Based on the information provided, the causes of the patient's anemia that cannot be excluded based on the findings of the history and physical examination and previous laboratory studies include:  Acute hemorrhage Chronic hemorrhage Anemia of chronic disease Intrinsic red blood cell abnormality Hemolysis Hypersplenism Vitamin B12 deficiency Myelodysplastic syndrome Chronic myeloid leukemia Note that more information and tests are likely needed to determine the cause of the patient's anemia. | Based on the findings of the history and physical examination, the causes of the patient's anemia that cannot be excluded at this point include but are not limited to:  Blood loss anemia: Despite the patient's denial of visible bleeding, other sources of blood loss such as gastrointestinal bleeding, heavy menstrual bleeding or cervical/uterine bleeding cannot be ruled out based on the history and examination. Vitamin deficiency anemia: Although the patient is a vegetarian, it is possible that she has a deficiency in vitamin B12 or folate, which are crucial for the production of red blood cells. Hemolytic anemia: The patient's history of a low-grade fever, diffuse joint pain, and rash 2 months prior raises a possibility of autoimmune or infectious causes of hemolytic anemia. Bone marrow disorder: Further evaluation, such as a bone marrow biopsy, is needed to rule out causes such as myelodysplastic syndrome, aplastic anemia, or other hematologic disorders. These potential causes will require further evaluation, such as laboratory tests and imaging studies, to determine the underlying cause of the patient's anemia. |
| M2_DIAG | It is not possible to determine which studies are most appropriate without additional information regarding the patient's symptoms, medical history, and the reason for obtaining these tests. A doctor or healthcare professional would be the best person to provide this information. | Complete blood count with differential Peripheral blood smear Reticulocyte count Serum iron and iron-binding capacity Serum C-reactive protein level Serum direct antiglobulin test (Coombs test) | Given the limited information provided, the most appropriate studies would be:  Complete blood count with differential Peripheral blood smear Serum iron and iron-binding capacity Serum C-reactive protein level Chest and abdominal CT scan |
| M2_DX | Pure red cell aplasia and Intrinsic red blood cell abnormality are the differential diagnoses that best account for the patient's findings. The peripheral smear showed numerous spherocytes and a low reticulocyte count, which are indicative of a red blood cell disorder. Additionally, the low hemoglobin and hematocrit levels are consistent with a red blood cell abnormality. The normal appearance of white blood cells and platelets on the peripheral smear, along with normal electrolyte levels and liver function tests, suggest that the cause of the anemia is not due to hypersplenism or a myelodysplastic syndrome. The negative direct antiglobulin test rules out hemolysis. | Based on the patient's findings, the differential diagnoses that best account for the patient's condition are intrinsic red blood cell abnormality and hemolysis. The peripheral smear findings of numerous spherocytes suggest that there may be a problem with the red blood cell membrane, which could be intrinsic or due to an autoimmune process. The low reticulocyte count, low serum iron, and elevated LDH (indicator of hemolysis) support the presence of hemolysis. | Based on the results of the tests, the differential diagnoses that best account for the patient's findings are Hemolysis and Hypersplenism. The peripheral blood smear showed numerous spherocytes, which is consistent with hemolysis. The low reticulocyte count, a laboratory measurement of the bone marrow's response to anemia, and the elevated total bilirubin levels, a byproduct of red blood cell breakdown, also support the diagnosis of hemolysis. Hypersplenism is suggested by the low hemoglobin and hematocrit levels, the low red blood cell count, and the low platelet count. |
| M2_DIAG | Bone marrow biopsy and PCR testing for parvovirus B19 infection. | Bone marrow biopsy and PCR testing for parvovirus B19 infection. | Bone marrow biopsy, PCR testing for parvovirus B19 infection, and Hemoglobin electrophoresis. |
| M2_DX | The most likely diagnosis is Transient pure red cell aplasia triggered by parvovirus B19 infection. | The most likely diagnosis at this time is Transient pure red cell aplasia triggered by parvovirus B19 infection. | The most likely diagnosis at this time is transient pure red cell aplasia triggered by parvovirus B19 infection. |
| M2_MANG | Intravenous immune globulin (IVIG) 1 gm/kg IV once per day for 2 days Monitor reticulocyte levels Repeat bone marrow examination at 2 weeks | Intravenous immune globulin (IVIG) 1 gm/kg IV once per day for 2 days Monitor reticulocyte levels Repeat bone marrow examination at 2 weeks | Transfuse 2 units packed RBCs Ferrous sulfate 325 mg orally twice a day Repeat bone marrow examination at 2 weeks Epoetin alfa 40,000 units subcutaneously Monitor reticulocyte levels |
| M2_Q1 | Acute hemorrhage, Chronic disease, Iron deficiency, Vitamin B12 deficiency | Acute hemorrhage Chronic disease (anemia of chronic disease) Iron deficiency Vitamin B12 deficiency | Acute hemorrhage, chronic disease, iron deficiency, and Vitamin B12 deficiency can cause normocytic anemia. |
| M2_Q2 | Hemolytic anemia | Hemolytic anemia and iron deficiency anemia typically manifest as increased reticulocyte count. | Hemolytic anemia and Iron deficiency anemia typically manifest as increased reticulocyte count. |
| M2_Q3 | Indications for blood transfusion include:  Hemoglobin <7.0 gm/dL Hemoglobin between 7.0 and 10.0 gm/dL with symptoms such as chest pain and exertional dyspnea Hemoglobin 9.0 gm/dL with resting dyspnea | The following are indications for blood transfusion:  Hemoglobin <7.0 g/dL Hemoglobin 8.0 g/dL with chest pain and exertional dyspnea Hemoglobin 9.0 g/dL with resting dyspnea The decision to transfuse should be based on the patient's symptoms, overall clinical condition, and laboratory values, and not just on a single hemoglobin value. In general, transfusions are usually not recommended for hemoglobin levels between 7.0 and 10.0 g/dL unless the patient has symptoms such as chest pain, exertional dyspnea or resting dyspnea. | Hemoglobin <7.0 gm/dL Hemoglobin 8.0 gm/dL with chest pain and exertional dyspnea Hemoglobin 9.0 gm/dL with resting dyspnea |
| M3_DIFF | At this time, the following differential diagnoses cannot be excluded: Ankylosing spondylitis, Chronic back injury, Idiopathic scoliosis. | Based on the history and physical examination, the following differential diagnoses cannot be excluded:  Chronic back injury Idiopathic scoliosis The patient has reported intermittent upper back pain and stiffness with mild paraspinal tenderness and asymmetrical scapulae noted on examination. Further investigation such as imaging and specialist referral may be necessary to determine the exact diagnosis and rule out other conditions. | At this time, the following differential diagnoses cannot be excluded: Ankylosing spondylitis, Chronic back injury, Idiopathic scoliosis. |
| M3_DIAG | Based on the information provided in the case, the most appropriate next steps would be:  X-ray of the cervical, thoracic, and lumbar spine: This will help to visualize the spine and determine if there is any structural abnormality that could be causing the back pain and stiffness. Evaluate for joint hypermobility: The presence of joint hypermobility can indicate the presence of Ehlers-Danlos syndrome, a condition that can cause back pain and other musculoskeletal symptoms. Determine the ratio of upper and lower body segments: This will help to determine if there is any structural abnormality in the patient's body that could be contributing to the back pain and stiffness. Determine the ratio of wingspan to height: This will help to determine if Marfan syndrome, a condition that can cause back pain and other musculoskeletal symptoms, is present. | Based on the examination results, the following next steps are appropriate:  Evaluate scoliosis further with MRI of the spine to determine the extent and cause. Consider additional tests to assess for underlying conditions that may be causing the scoliosis, such as neurological or connective tissue disorders. | Evaluate for joint hypermobility, do finger-nose test, do straight-leg raising test. |
| M3_DX | Based on the results, the most likely diagnosis is Adolescent idiopathic scoliosis. | Adolescent idiopathic scoliosis is the most likely diagnosis. | Adolescent idiopathic scoliosis |
| M3_MANG | Based on the information provided, the most appropriate management steps for this patient are:  Ibuprofen 400 mg orally every 8 hours as needed for back discomfort Back x-rays and examination every 6 months to monitor scoliosis angle | Based on the results, the most appropriate management steps for this patient are:  Ibuprofen 400 mg orally every 8 hours as needed for back and discomfort Back x-rays and examination every 6 months to monitor scoliosis angle | Back x-rays and examination every 6 months to monitor scoliosis angle Ibuprofen 400 mg orally every 8 hours as needed for back and discomfort |
| M3_Q1 | 10° | 10° | 10° |
| M3_Q2 | His genital exam being consistent with Tanner Stage III and not yet shaving are physical examination findings that indicate the patient has not yet completed skeletal growth. | His genital exam consistent with Tanner Stage III. | His genital exam is consistent with Tanner Stage III. |
| M3_Q3 | Age 10 years | Age 10 years | Age 10 years |
| M3_Q4 | The goal of treatment with bracing is to achieve a curve less than 50° at maturity. | The goal of treatment with bracing is to achieve a curve less than 20° of curvature at maturity. | 20° |
| M4_DIFF | At this time, the following differential diagnoses cannot be excluded based on the history and physical examination in this patient:  Non-ST-segment elevation myocardial infarction (NSTEMI)  Unstable angina  Acute aortic dissection  Pulmonary embolism (PE)  Acute pericarditis  Severe aortic stenosis (AS)  Stable angina  STEMI  These diagnoses should be considered due to the patient's symptoms of increasing exertional chest pain, radiation to the left arm, dyspnea, and nausea, which are all concerning for possible cardiac conditions. The patient's past medical history of hypertension, COPD, and osteoarthritis, as well as his smoking history, also support these differential diagnoses. | Acute aortic dissection, Non-ST-segment elevation myocardial infarction (NSTEMI), Pulmonary embolism (PE), Stable angina, Unstable angina cannot be excluded based on the history and physical examination in this patient. | Non-ST-segment elevation myocardial infarction (NSTEMI), Pulmonary embolism (PE), Stable angina, Unstable angina, Acute aortic dissection, Pneumonia, Psychological factors, Severe aortic stenosis (AS), ST-segment elevation myocardial infarction (STEMI) cannot be excluded based on the history and physical examination. |
| M4_DIAG | Arterial blood gas values (ABGs), Cardiac monitoring, ECG, Reassess patient 5 min after administering sublingual nitroglycerin, Serum troponin level. | Cardiac monitoring  ECG  Chest x-ray, posteroanterior (PA) and lateral  Serum troponin level  Reassess patient 5 min after administering sublingual nitroglycerin | Cardiac monitoring, ECG, Reassess patient 5 min after administering sublingual nitroglycerin. |
| M4_DX | The most likely diagnosis at this time is Non-ST-segment elevation myocardial infarction (NSTEMI). | The most likely diagnosis at this time is Non-ST-segment elevation myocardial infarction (NSTEMI) based on the elevated serum troponin level and the resolution of symptoms after administration of nitroglycerin. | The most likely diagnosis is Non-ST-segment elevation myocardial infarction (NSTEMI) |
| M4_MANG | Admit the patient to telemetry unit, consult cardiology, aspirin 81 mg po, and clopidogrel 75 mg po. | Admit the patient to telemetry unit, consult cardiology, Aspirin 325 mg po, Clopidogrel 75 mg po, Atorvastatin 80 mg po. | Admit the patient to telemetry unit, consult cardiology, aspirin 81 mg po, clopidogrel 75 mg po, heparin 60 IU/kg(max 4000 units) loading dose and continue infusion at 12 IU/kg/h. |
| M4_Q1 | COPD, diabetes, dyslipidemia, hypertension, and tobacco use are considered risk factors for coronary artery disease. | COPD, Diabetes, Dyslipidemia, Hypertension, and Tobacco use are considered risk factors for coronary artery disease. | COPD, Diabetes, Dyslipidemia, Hypertension, Tobacco use are considered risk factors for coronary artery disease. |
| M4_Q2 | Urgent cardiac catheterization in a NSTEMI presentation would warrant the following factors:  Continued chest pain despite administration of nitroglycerin and morphine Progression of ECG changes to ST elevations Serum troponin T level > 5 ng/ mL Ventricular tachyarrhythmias | Hypotension, Serum troponin T level > 5 ng/ mL, and Ventricular tachyarrhythmias | Continued chest pain despite administration of nitroglycerin and morphine Hypotension Progression of ECG changes to ST elevations Ventricular tachyarrhythmias |
| M4_Q3 | The TIMI score helps categorize risk level for:  Death from acute coronary syndrome within 14 days  Myocardial infarction | Death from acute coronary syndrome within 14 days  Myocardial infarction | Death from acute coronary syndrome within 14 days Myocardial infarction |
| M5_DIFF | Aortic dissection, Pulmonary embolism, and Community-acquired pneumonia are common and/or dangerous differential diagnoses that cannot be excluded based on the history and physical examination of this patient. | Aortic dissection, Pulmonary embolism, Community-acquired pneumonia. | Aortic dissection, Pulmonary embolism, and Community-acquired pneumonia are common and/or dangerous differential diagnoses that cannot be excluded based on the history and physical examination of this patient. |
| M5_DIAG1 | An Electrocardiogram (ECG), a chest x-ray, portable, and a serum troponin concentration are essential to do before the patient leaves the ED. The other tests, such as Arterial blood gas values, Chest x-ray, posteroanterior and lateral, CT scan of the chest, Serum D-dimer levels, Complete blood count (CBC), Pro-brain natriuretic peptide, and Complete metabolic panel (CMP), may also be done depending on the results of the initial tests and the clinician's discretion. | Chest x-ray, portable and Electrocardiogram (ECG) are essential before the patient leaves the ED. | Chest x-ray, portable, Electrocardiogram (ECG), and Serum troponin concentration are essential steps to be performed before the patient leaves the ED. The other tests could also be done if needed based on the results of the initial tests and the clinical presentation of the patient. |
| M5_DIAG2 | The most appropriate step to guide diagnosis and treatment would be a CT scan of the chest with IV contrast, based on the elevated serum D-dimer level. Pulmonary embolism is a serious condition and should be ruled out as a potential cause. Cardiac catheterization may also be considered as a diagnostic option but is typically a more invasive procedure than a CT scan. Pulmonary function testing and Doppler ultrasonography of the legs may not be as relevant in this case, but could be considered depending on the clinical presentation and other factors. | Further evaluation and diagnosis might require additional tests and assessments, such as CT scan of the chest with IV contrast, and may include:  Clinical evaluation of symptoms Cardiac biomarkers (e.g. serum troponin) Electrocardiogram (ECG) Cardiac imaging tests (e.g. echocardiogram, stress test, angiogram) D-dimer testing (for suspected pulmonary embolism) Additional blood tests (e.g. complete metabolic panel, pro-brain natriuretic peptide) The most appropriate steps would depend on the patient's individual clinical presentation, risk factors, and results of initial testing. A healthcare professional would be best equipped to make these determinations based on a full evaluation of the patient. | Based on the information provided, the most appropriate steps to guide diagnosis and treatment are CT scan of the chest with IV contrast and/or Doppler ultrasonography of the legs. These tests can help evaluate the presence of a potential pulmonary embolism, which could explain the elevated D-dimer levels. The elevated D-dimer levels along with the normal troponin I levels suggest the absence of acute myocardial infarction, but further testing may be required to rule out other potential causes of the elevated D-dimer levels. |
| M5_DX | Given the results of the laboratory testing and the CT scan of the chest with IV contrast, the most likely diagnosis at this time is Pulmonary embolism (PE). | The most likely diagnosis at this time is Pulmonary embolism (PE). | Pulmonary embolism (PE) |
| M5_MANG | Admit the patient to the hospital for further treatment, Heparin 80 U/kg IV bolus followed by 18 U/kg/h IV infusion, and Warfarin 5 mg po once/day at bedtime are the most appropriate treatments. | Admit the patient to the hospital for further treatment, Heparin 80 U/kg IV bolus followed by 18 U/kg/h IV infusion, and Rivaroxaban 15 mg po bid. | Admit the patient to the hospital for further treatment, Heparin 80 U/kg IV bolus followed by 18 U/kg/h IV infusion, Warfarin 5 mg po once/day at bedtime, Rivaroxaban 15 mg po bid. |
| M5_Q1 | Hypotension and Tachycardia are the two most common vital sign abnormalities in patients with acute pulmonary embolism. | Hypotension, Tachycardia | Hypotension, Tachycardia. |
| M5_Q2 | Dabigatran and rivaroxaban are approved for outpatient treatment of pulmonary embolism. | Enoxaparin, Dabigatran, and Fondaparinux are approved for outpatient treatment of pulmonary embolism. Aspirin is not approved for this use. | Dabigatran and Enoxaparin are approved for outpatient treatment of pulmonary embolism. |
| M5_Q3 | The components of the Wells Criteria for pulmonary embolism include:  Heart rate > 100 beats/min Immobilization for at least 3 days or history of surgery in the previous 4 weeks History of cancer (with treatment within the past 6 months or currently active) Hemoptysis Previous deep vein thrombosis or pulmonary embolism Alternative diagnosis is less likely than pulmonary embolism. | Heart rate > 100 beats/min and immobilization for at least 3 days or history of surgery in the previous 4 weeks. | Heart rate > 100 beats/min Immobilization for at least 3 days or history of surgery in the previous 4 weeks |
| M6_DIFF | Acute coronary syndrome (ACS), unstable angina, or acute MI  Pulmonary embolism (PE) | Based on the history and physical examination findings, the following differential diagnoses cannot be excluded at this time:  Acute coronary syndrome (ACS), unstable angina, or acute MI Stable angina Musculoskeletal pain syndrome, including costochondritis Gastroesophageal reflux disease (GERD) Pericarditis. | Acute coronary syndrome (ACS), unstable angina, or acute MI, Aortic dissection, Pericarditis, Pulmonary embolism (PE), and Stable angina cannot be excluded at this time. |
| M6_DIAG1 | ECG, serum D-dimer level | The following steps are essential to do before the patient leaves the clinic:  ECG Pulse oximetry. | ECG and possibly a Chest x-ray would be essential to do before the patient leaves the clinic to help further evaluate the cause of the chest pain. The need for a pulse oximetry and serum D-dimer level would depend on other factors and the healthcare provider's clinical judgment. |
| M6_DIAG2 | Serum troponin level | Serum troponin level and an ECG would be the most appropriate steps to guide diagnosis and treatment based on the previous findings. Elevated troponin levels can indicate cardiac injury and ECG can help to detect any underlying cardiac arrhythmias or ischemic changes that could be contributing to the patient's symptoms. Other tests such as cardiac catheterization, CT angiography, echocardiography, electron beam CT for calcium score, treadmill stress test with ECG, and treadmill stress test with nuclear imaging could be considered based on the results of the initial testing and the patient's clinical condition. | The most appropriate step to guide diagnosis and treatment would be to check a serum troponin level, which is a sensitive and specific biomarker for myocardial injury and can help determine the likelihood of an acute coronary syndrome. Other tests, such as a treadmill stress test with ECG or a CT angiography of the chest, may also be considered based on the healthcare provider's clinical judgment and assessment of the patient's symptoms and risk factors. |
| M6_DX | Acute coronary syndrome | Acute coronary syndrome | Acute coronary syndrome. |
| M6_MANG | Refer to a cardiologist for cardiac catheterization, Admit to the hospital telemetry unit, Reinforce need to adhere to antihypertensive treatment, Aspirin 81 mg once/day, Nitroglycerin 0.4 mg sublingual as needed for chest pain | Admit to the hospital telemetry unit, Refer to a cardiologist for cardiac catheterization, Aspirin 81 mg once/day, Nitroglycerin 0.4 mg sublingual as needed for chest pain | Refer to a cardiologist for cardiac catheterization, Admit to the hospital telemetry unit, Aspirin 325 mg once/day, Metoprolol 25 mg twice daily, Nitroglycerin 0.4 mg sublingual as needed for chest pain. |
| M6_Q1 | Chest pain characteristics that suggest cardiac rather than noncardiac chest pain:  Substernal location Pressure-like quality Triggered by exertion Relieved by nitroglycerin Lasting longer than a few seconds. | Chest pain characteristics that suggest cardiac rather than noncardiac chest pain include:  Substernal location Pressure-like quality Triggered by exertion Relieved by nitroglycerin Lasting more than a few seconds | Chest pain characteristics that suggest cardiac chest pain include:  Pressure-like quality Substernal location Triggered by exertion Relieved by nitroglycerin These symptoms are typical of angina, which is chest pain caused by reduced blood flow to the heart due to blockages in the coronary arteries.\ |
| M6_Q2 | Chronic obstructive pulmonary disease (COPD) Atrial fibrillation Uncontrolled hypertension | The following patient characteristics are relative contraindications to dobutamine stress echocardiography:  Aortic stenosis, severe Atrial fibrillation Hypertension, uncontrolled | The following patient characteristics are relative contraindications to dobutamine stress echocardiography:  Aortic stenosis, severe Atrial fibrillation Chronic obstructive pulmonary disease (COPD) Hypertension, uncontrolled |
| M6_Q3 | Pain occurring at rest and increasing in frequency or severity. | Pain increasing in duration, pain occurring at rest, and pain occurring more than 5 times/day would have suggested unstable angina and therefore would have required a more urgent workup. | Pain occurring at rest and pain increasing in severity suggest unstable angina and would have required a more urgent workup. |
| M6_Q4 | Calcium score | Calcium score and stress ECG are appropriate for further risk stratification of this patient. | Calcium score and stress ECG would be appropriate for further risk stratification of this patient. |
| M7_DIFF | The differential diagnoses that should not be excluded are:  Alzheimer disease Chronic bilateral subdural hematoma Heart failure Parkinson disease Stroke | The 68-yr-old woman presents with 4-mo history of gradually increasing lethargy, worsening memory, and changes in her appearance, voice, and respiratory system. She feels tired all the time, has a poor appetite but gained 5 kg, has dry skin, hoarse voice, fullness in throat, exertional shortness of breath, leg swelling, and depression. The patient has hypertension, atrial fibrillation, and takes multiple medications. Physical examination shows periorbital edema, enlarged thyroid, fine crackles, irregular heart rate, leg edema, and mild discomfort in the left iliac fossa. She has a positive Tinel's sign on the right side. | Based on the history and physical examination findings, the 68-year-old woman presents with symptoms of lethargy, worsening memory, and decreased mobility, as well as signs of edema, dry skin and hair, hoarseness, and a swollen thyroid gland. The patient has a history of hypertension, atrial fibrillation, and appendectomy. She has been taking multiple medications to control her hypertension and AF. The physical examination also reveals irregular heart rate, mild edema of the lower extremities, decreased deep tendon reflexes, and proximal weakness of the lower extremities. These symptoms suggest possible underlying medical conditions, such as thyroid disease, cardiovascular disease, or a neurodegenerative condition. Further evaluation, including laboratory tests, imaging studies, and neurologic examination, is needed to determine the cause of her symptoms and develop an appropriate treatment plan. |
| M7_DIAG1 | Complete blood count (CBC), Electrocardiogram (ECG), Serum electrolyte, BUN, and creatinine concentrations, Thyroid function tests. | Serum electrolyte, BUN, and creatinine concentrations Urinalysis Complete blood count (CBC) | It is not possible to determine the most appropriate tests without additional information about the patient's symptoms, medical history, and reasons for the examination. However, some common tests for a general evaluation may include:  Complete blood count (CBC) Serum electrolyte, BUN, and creatinine concentrations Urinalysis Electrocardiogram (ECG) Chest x-ray |
| M7_DX1 | Hypothyroidism | Hypothyroidism | Hypothyroidism |
| M7_DIAG2 | Serum thyroid peroxidase (TPO) antibodies, Thyroid ultrasound | Serum thyroid peroxidase (TPO) antibodies, Thyroid ultrasound | Serum TPO antibodies Thyroid ultrasound |
| M7_DX2 | Given the results of recent testing, the most likely diagnosis at this time is Hashimoto thyroiditis. | Hashimoto thyroiditis | The most likely diagnosis based on the results of the serum thyroid peroxidase (TPO) antibodies test is Hashimoto thyroiditis. |
| M7_MANG | L-thyroxine 25 mcg po once/day | L-thyroxine 25 mcg po once/day | L-thyroxine 25 mcg po once/day |
| M7_Q1 | Hashimoto thyroiditis, Iodine deficiency, Radioiodine administration, and Radiotherapy to the neck can all cause primary hypothyroidism. | Hashimoto thyroiditis, iodine deficiency, radioiodine administration, radiotherapy to the neck, postpartum thyroiditis, and drug-induced thyroiditis can cause primary hypothyroidism. | Hashimoto thyroiditis, iodine deficiency, radioiodine administration, postpartum thyroiditis, and radiotherapy to the neck can cause primary hypothyroidism. |
| M7_Q2 | Addison's disease, Celiac disease, Pernicious anemia, Vitiligo, and Polyglandular autoimmune syndrome are associated with autoimmune hypothyroidism. | Hashimoto thyroiditis, autoimmune polygendocrine syndrome, and vitiligo are associated with autoimmune hypothyroidism. | Autoimmune hypothyroidism is associated with the following conditions:  Hashimoto thyroiditis Polyglandular autoimmune syndrome Vitiligo |
| M7_Q3 | Amiodarone, Carbamazepine, Lithium, Phenytoin, and Rifampin. | Amiodarone, Lithium, Phenytoin, Carbamazepine, Interferon alpha, Rifampin. | Amiodarone, Lithium, Phenytoin, Carbamazepine, Interferon alpha, Rifampin, and Sunitinib can require reassessment of thyroid function and may require an adjustment of the L-thyroxine dose.     Michael Pang What are the possible complications of hypothyroidism? (You may select more than one option.)    Acute heart failure    Diabetes    Encephalopathy    Hypercalcemia    Hyperkalemia    Hyponatremia    Normocytic anemia    Megacolon    Myxedema coma   Myxedema coma Acute heart failure Normocytic anemia Encephalopathy Hyponatremia |
| M7_Q4 | The possible complications of hypothyroidism include:  Myxedema coma Acute heart failure Normocytic anemia Encephalopathy | Acute heart failure  Encephalopathy  Myxedema coma  Normocytic anemia | Myxedema coma Acute heart failure Normocytic anemia Encephalopathy Hyponatremia |
| M8_DIFF | The following differential diagnoses cannot be excluded based on the history and physical examination in this patient:  Alcohol intoxication Alcohol withdrawal Metabolic encephalopathy Dementia | Acute stroke, alcohol intoxication, alcohol withdrawal, metabolic encephalopathy, seizures | Acute stroke, alcohol intoxication, alcohol withdrawal, drug encephalopathy, metabolic encephalopathy, and seizures are common and/or dangerous differential diagnoses that cannot be excluded based on the history and physical examination of this patient. |
| M8_DIAG1 | Given the previous findings, the following are the most appropriate steps to guide diagnosis and treatment:  Serum alcohol level Complete metabolic profile Electroencephalography (EEG) CT scan of the brain without contrast Urine toxicology screen | CT scan of the brain without contrast, Complete metabolic profile, Electroencephalography (EEG), Serum alcohol level, Urine toxicology screen. | The most appropriate steps to guide diagnosis and treatment would include a CT scan of the brain without contrast, serum alcohol level, and a urine toxicology screen. Additionally, a complete metabolic profile and serum vitamin B12 level may also be considered to further evaluate for potential metabolic causes of confusion. |
| M8_DIAG2 | Based on the CT scan findings, the most appropriate tests to guide diagnosis and treatment would be:  MRI with and without contrast of the head Lumbar puncture | MRI with and without contrast of the head | MRI with and without contrast of the head and lumbar puncture are the most appropriate tests to guide diagnosis and treatment based on the CT scan findings. These tests can help to further evaluate the nature of the ill-defined rounded mass and rule out any intracranial pathology. |
| M8_DX | Seizure due to meningioma | Seizure due to meningioma | Seizure due to meningioma |
| M8_MANG | Admit the patient to the hospital for surgical resection of the meningioma | Admit the patient to the hospital for surgical resection of the meningioma, Discharge the patient with follow-up with a neurologist if symptoms recur. | The most appropriate next steps would be to admit the patient to the hospital for surgical resection of the meningioma, and to discharge the patient with follow-up with a neurologist if symptoms recur. |
| M8_Q1 | Haloperidol and penicillin have been reported to induce seizures. | Seizure-inducing drugs:  Haloperidol Aminophylline Penicillin Finasteride Mycophenalate mofetil | Haloperidol, Aminophylline, Prednisone and Mycophenalate mofetil have been reported to induce seizures. |
| M8_Q2 | Lamotrigine and valproate are most commonly associated with Stevens-Johnson syndrome in adults. | Lamotrigine is most commonly associated with Stevens-Johnson syndrome in adults. | Lamotrigine and valproate are most commonly associated with Stevens-Johnson syndrome in adults. |
| M8_Q3 | Common side effects of anticonvulsant drugs include:  Fatigue Drowsiness Nausea Dizziness Diarrhea Rash Insomnia Headache Weight changes Tremors Cognitive/memory impairment Blurred vision Tinnitus Nervousness/anxiety Tremors Vomiting. | Fatigue, insomnia, nausea, vomiting, diarrhea, impotence, and rash are common side effects of anticonvulsant drugs. | Common side effects of anticonvulsant drugs may include:  Fatigue Dizziness Nausea Vomiting Diarrhea Insomnia Rash Tinnitus Weight gain Tremors Drowsiness Impaired coordination or balance |
| M9_DIFF | Acute bronchitis, asthma, coronary ischemia, gastroesophageal reflux disease (GERD), medication-induced cough, pneumonia, postviral cough, sarcoidosis, and tuberculosis (TB) cannot be excluded based on history and physical examination in this patient. | Based on the patient's history and physical examination, the differential diagnoses that cannot be excluded at this time include:  Acute bronchitis Asthma Lung cancer Medication-induced cough Pneumonia Sarcoidosis Tuberculosis (TB) It is important to note that this list is not exhaustive and other conditions could also be considered. Further evaluation and testing may be necessary to confirm the diagnosis. | Based on the history and physical examination, the following differential diagnoses cannot be excluded:  Acute bronchitis Asthma Gastroesophageal reflux disease (GERD) Medication-induced cough Pneumonia Postviral cough Sarcoidosis Tuberculosis (TB) |
| M9_DIAG | Chest x-ray and pulmonary function testing are the most appropriate next steps. | Based on the patient's symptoms of cough, wheezing, and chest pain, the most appropriate next step would be:  Pulmonary function testing This would help determine if the patient has an underlying respiratory condition such as asthma or bronchitis and evaluate the severity of their symptoms. Other tests, such as a chest x-ray or CT scan, may also be considered if the results of pulmonary function testing suggest the need for further evaluation. | Based on the history and physical examination, the most appropriate next step would be:  Pulmonary function testing |
| M9_MANG | Albuterol inhaler 2 puffs q 6 h and prn for wheezing, Budesonide inhaler 1 puff bid, Combined ipratropium/albuterol inhaler 2 puffs q 6 h, Prednisone 40 mg po once/day for 5 days. | Albuterol inhaler 2 puffs q 6 h and prn for wheezing  Budesonide inhaler 1 puff bid  Combined ipratropium/albuterol inhaler 2 puffs q 6 h  Omeprazole 20 mg po once/day | Albuterol inhaler 2 puffs q 6 h and prn for wheezing Budesonide inhaler 1 puff bid Combined ipratropium/albuterol inhaler 2 puffs q 6 h Omeprazole 20 mg po once/day |
| M9_Q1 | Influenza vaccine Pneumococcal polysaccharide vaccine (PPSV23) | Influenza vaccine and Pneumococcal polysaccharide vaccine (PPSV23) are particularly important for this patient to receive on a yearly basis. | Influenza vaccine and Pneumococcal polysaccharide vaccine (PPSV23) |
| M9_Q2 | HIV infection, working in a day-care center, use of tumor necrosis factor-alpha inhibitors. | The following are risk factors for tuberculosis:  HIV infection Use of tumor necrosis factor-alpha inhibitors Working in a day-care center Living in a crowded or high-density housing Substance abuse (including injection drug use) Silicosis (a lung disease caused by inhaling silica dust) Tobacco smoking Low body weight Alcoholism Diabetes Chronic kidney disease Malnutrition Immune-suppressing conditions (such as cancer, organ transplant, or treatment with immunosuppressive drugs) | HIV infection Use of tumor necrosis factor-alpha inhibitors |
| M9_Q3 | Teaching the patient how to use a peak flow meter at home | Teaching the patient how to use a peak flow meter at home. | Teaching the patient how to use a peak flow meter at home |
| M10_DIFF | Based on the history and physical examination, the common and/or dangerous differential diagnoses that cannot be excluded are:  Asthma exacerbation Bronchiolitis Foreign body aspiration Pneumonia | Allergic reaction, asthma exacerbation, bronchiolitis, foreign body aspiration, pneumonia. | Asthma exacerbation, Bronchiolitis, Foreign body aspiration, and Pneumonia. |
| M10_DIAG | Based on the information provided, the most appropriate next steps are:  Albuterol nebulizer Chest x-ray (Anteroposterior (AP), lateral, and expiratory views) Pulse oximetry Complete blood count (CBC) with differential. | Chest x-ray: Anteroposterior (AP), lateral, and expiratory views, pulse oximetry, albuterol nebulizer. | Albuterol nebulizer, Chest x-ray (Anteroposterior (AP), lateral, and expiratory views), Complete blood count (CBC) with differential, and Pulse oximetry. |
| M10_MANG1 | Based on the results of the pulse oximetry and chest x-ray, the most appropriate next step would be to:  Admit the child for observation Repeat attempt at chest x-ray expiratory view. Given the child's difficulty breathing and low oxygen saturation, admission for observation and further evaluation is warranted. The lack of a clear expiratory view on the chest x-ray suggests there may be an underlying issue that requires further imaging. | Admit the child for observation, repeat attempt at chest x-ray expiratory view. | Admit the child for observation and Repeat attempt at chest x-ray expiratory view. |
| M10_DX | Based on the history and physical examination and the normal anteroposterior and lateral chest x-ray views, the most likely diagnosis at this time is asthma with air trapping. However, the limited information provided does not allow for a definite conclusion, and further testing may be needed to rule out other possible diagnoses. | Asthma with air trapping | Without further information, it is not possible to determine the most likely diagnosis based on the information provided. Additional testing and observation may be needed to arrive at a diagnosis. |
| M10_MANG2 | Consult ENT or pulmonary specialist for bronchoscopy and suction airway with a flexible catheter. | Consult ENT or pulmonary specialist for bronchoscopy. Suction airway with a flexible catheter. | At this time, the most appropriate treatment orders would be:  Consult ENT or pulmonary specialist for bronchoscopy Sputum Gram stain and culture Amoxicillin/clavulanate 45 mg/kg IV q 12 h |
| M10_Q1 | Cough, drooling, unilateral diminished breath sounds, and wheezing are considered typical manifestations of acute bronchial foreign body in children. | Cough, drooling, unilateral diminished breath sounds, and wheezing. | Cough, wheezing, unilateral diminished breath sounds, and drooling are considered typical manifestations of acute bronchial foreign body in children. |
| M10_Q2 | The typical complications of an unrecognized bronchial foreign body include:  Atelectasis Bronchiectasis Lung abscess Pneumonia | Cough, wheezing, and unilateral diminished breath sounds are typical manifestations of acute bronchial foreign body in children.  Bronchiectasis, lung abscess, and pneumonia are typical complications of an unrecognized bronchial foreign body. | The typical complications of an unrecognized bronchial foreign body are:  Atelectasis Bronchiectasis Lung abscess Pneumonia |
| M10_Q3 | The most common location of foreign bodies aspirated by young children is the bronchi. | The most common location of foreign bodies aspirated by young children is the bronchi. | The most common location of foreign bodies aspirated by young children is the bronchi. |
| M10_Q4 | Right | In a child aged 1 year to 3 years old, a foreign body is most likely to be located in the right bronchus. | In a child aged 1 year to 3 years old, a foreign body is most likely to be located in the right bronchus. |
| M11_DIFF | Based on the history and physical examination, the following differential diagnoses cannot be excluded in this patient:  Acute bronchitis COPD exacerbation Lung cancer New-onset heart failure Pneumonia Pleural effusion. | Based on the information provided, the differential diagnoses that cannot be excluded based on history and physical examination in this patient are:  Acute bronchitis COPD exacerbation Pneumonia Pulmonary embolism (PE) TB | Based on the history and physical examination in this patient, the differential diagnoses that cannot be excluded at this time are: Acute bronchitis, pneumonia, COPD exacerbation, lung cancer, and pulmonary embolism (PE). |
| M11_DIAG | Chest x-ray, chest CT scan, arterial blood gasses (ABGs), pulse oximetry, ECG, complete blood count (CBC), blood and sputum cultures, serum chemistry profile, serum D-dimer level, serum troponin level. | Chest x-ray, pulse oximetry, blood and sputum cultures, and possibly a chest CT scan. | Chest x-ray, Chest CT scan, Pulse oximetry, Arterial blood gasses (ABGs), ECG, Serum troponin level, Blood and sputum cultures. |
| M11_DX | Community-acquired pneumonia | Given the information available, the most likely diagnosis would be community-acquired pneumonia. | Community-acquired pneumonia |
| M11_MANG | Azithromycin 500 mg po once/day, then 250 mg po once/day Oxygen 4 L/min by nasal cannula | Azithromycin 500 mg po once/day, then 250 mg po once/day Oxygen 4 L/min by nasal cannula Schedule follow-up visit in 6 wk for repeat chest x-ray | Azithromycin 500 mg po once/day, then 250 mg po once/day and Oxygen 4 L/min by nasal cannula |
| M11_Q1 | Streptococcus pneumonia, Haemophilus influenzae, and Mycoplasma pneumoniae are the most likely causes of community-acquired pneumonia in this patient. | The most likely cause of community-acquired pneumonia in this patient is Streptococcus pneumonia. | Streptococcus pneumonia, Haemophilus influenzae, and Mycoplasma pneumoniae are the most likely causes of community-acquired pneumonia in this patient. |
| M11_Q2 | History of cancer, recent immobilization for 3 days, and visible leg swelling would increase the patient's pre-test probability of pulmonary embolism (PE). | The following factors, if present, would increase the patient's pre-test probability of pulmonary embolism (PE):  Exertional dyspnea Heart rate: 110 beats/min Pleuritic chest pain Recent immobilization for 3 days Visible leg swelling | History of cancer, recent immobilization for 3 days, pleuritic chest pain, and visible leg swelling would increase the patient's pre-test probability of pulmonary embolism (PE). |
| M11_Q3 | Altered mental status Oxygen saturation: 92% Temperature: 40º C | The following factors, if present, would suggest increased risk of a poor outcome in patients with community-acquired pneumonia:  Altered mental status Oxygen saturation: 92% Temperature: 40º C | Altered mental status, oxygen saturation of 92%, and temperature of 40º C suggest increased risk of a poor outcome in patients with community-acquired pneumonia. |
| M12_DIFF | Based on the history and physical examination, the following differential diagnoses cannot be excluded:  Pneumonia Pulmonary embolism Spontaneous pneumothorax Tension pneumothorax Thoracic aortic dissection | Asthma exacerbation, lung cancer, pneumonia, pulmonary embolism, spontaneous pneumothorax, and tension pneumothorax cannot be excluded based on the patient's history and physical examination. | Based on the history and physical examination, the following differential diagnoses cannot be excluded:  Asthma exacerbation Lung cancer Musculoskeletal back pain Pneumonia Pulmonary embolism Spontaneous pneumothorax Tension pneumothorax Thoracic aortic dissection |
| M12_DIAG | Given the patient's current history and examination findings, the following steps are most appropriate to do at this time:  Chest x-ray ECG D-dimer testing Oxygen 6 L by nasal cannula It is also likely that a CT angiography (CTA) of the chest or thoracic spine x-rays may be performed later in the evaluation, if indicated by the results of the initial tests. | Chest x-ray, oxygen 6 L by nasal cannula, and ECG are the most appropriate steps to do at this time based on the patient's history and examination findings. | Based on the patient's history and examination findings, the following steps are most appropriate to do at this time:  Chest x-ray ECG Oxygen 6 L by nasal cannula D-dimer testing A CT angiography (CTA) of the chest or thoracic spine x-rays may be considered if the patient's condition worsens or if the results of initial tests are not conclusive. |
| M12_MANG | Given the patient's current history and examination findings, the following steps are essential to do immediately:  Continue oxygen 6 L by nasal cannula Admit the patient to the hospital Other steps such as a CT scan of the chest with or without IV contrast, increasing supplemental oxygen, or chest tube placement may be necessary later in the evaluation and management, but these decisions should be made based on the results of initial tests and the patient's response to treatment. Discharging the patient, administering azithromycin, or scheduling follow-up chest x-ray in 1 day are not appropriate based on the limited information provided. Needle decompression, video-assisted thoracoscopic surgery, and chemical pleurodesis are more advanced management options that may be considered later in the evaluation, but are not essential at this time. | Continue oxygen 6 L by nasal cannula, and admit the patient to the hospital are essential steps to do immediately based on the patient's history and examination findings. | Based on the patient's history and examination findings, the following steps are essential to do immediately:  Continue oxygen 6 L by nasal cannula Increase supplemental oxygen to 100% via nonrebreathing face mask Admit the patient to the hospital CT scan of the chest with IV contrast The other options listed (administering azithromycin, discharging the patient, needle decompression, chest tube placement, and scheduling video-assisted thoracoscopic surgery and chemical pleurodesis) may be considered based on the results of the patient's imaging studies and the progression of their condition. |
| M12_Q1 | Approximately 20% of pneumothorax patients have primary spontaneous pneumothorax. | 20% | Approximately 20% of pneumothorax cases are classified as primary spontaneous pneumothorax. |
| M12_Q2 | 20s | 20s | The peak age of patients who develop primary spontaneous pneumothorax is in the late teens to early 30s. |
| M12_Q3 | Male sex, smoking, and family history are risk factors for primary spontaneous pneumothorax. | Male sex, Smoking, Family history. | Male sex and smoking are risk factors for primary spontaneous pneumothorax. |
| M13_DIFF | Acute decompensated heart failure (ADHF), Anemia, Aortic stenosis, Pulmonary embolism (PE), Volume overload from end-stage renal disease. | Acute decompensated heart failure (ADHF) Aortic stenosis Coronary artery disease (CAD) Mitral regurgitation Pulmonary embolism (PE) Volume overload from end-stage renal disease | Acute decompensated heart failure (ADHF) Anemia Aortic stenosis Chronic obstructive pulmonary disease (COPD) Coronary artery disease (CAD) Pulmonary embolism (PE) Volume overload from end-stage renal disease |
| M13_DIAG | Chest CT scan Chest x-ray (CXR) Electrocardiogram (ECG) Serum troponin I level Transthoracic echocardiography (TTE) | Based on the information in the history and physical examination, the most appropriate next steps to guide diagnosis and treatment are:  Electrocardiogram (ECG) Chest x-ray (CXR) Serum brain natriuretic peptide (BNP) level Transthoracic echocardiography (TTE) | Chest x-ray (CXR) Complete blood count (CBC) Electrocardiogram (ECG) Serum brain natriuretic peptide (BNP) level Transthoracic echocardiography (TTE) |
| M13_DX | Severe aortic stenosis | Severe aortic stenosis. | Severe aortic stenosis |
| M13_MANG | Admit to the hospital for emergency aortic valve surgery, Refer to cardiology to evaluate for surgery. | Admit to the hospital for emergency aortic valve surgery, Refer to cardiology to evaluate for surgery | Admit to the hospital for emergency aortic valve surgery, Refer to cardiology to evaluate for surgery. |
| M13_Q1 | Delayed carotid upstrokes, diminished or absent S2, displaced point of maximal impulse (PMI), early peaking systolic murmur, increase in murmur intensity with inspiration, and murmur that radiates to the carotid arteries. | The following physical examination findings suggest severe aortic stenosis:  Early peaking systolic murmur Increase in murmur intensity with inspiration Loud S2 Displaced point of maximal impulse (PMI) Murmur that radiates to the carotid arteries. | The physical examination findings that suggest severe aortic stenosis include:  Early peaking systolic murmur Increase in murmur intensity with inspiration Displaced point of maximal impulse (PMI) Murmur that radiates to the carotid arteries |
| M13_Q2 | 75-year-old man with 3-month history of exertional chest pressure and severe aortic stenosis on TTE, and 64-year-old asymptomatic woman with severe aortic stenosis and left ventricular ejection fraction (LVEF) of 40% meet criteria for aortic valve replacement. | Patients who meet criteria for aortic valve replacement include:  75-year-old man with 3-month history of exertional chest pressure and severe aortic stenosis on TTE 64-year-old asymptomatic woman with severe aortic stenosis and left ventricular ejection fraction (LVEF) of 40% | 75-year-old man with 3-month history of exertional chest pressure and severe aortic stenosis on TTE 64-year-old asymptomatic woman with severe aortic stenosis and left ventricular ejection fraction (LVEF) of 40% |
| M14_DIFF | At this time, the following differential diagnoses cannot be excluded based on history and physical examination in this patient: Atrial fibrillation, Atrial flutter, Atrioventricular nodal reentrant tachycardia (AVNRT), Premature atrial complexes (PACs), Premature ventricular complexes (PVCs), Nonsustained ventricular tachycardia (NSVT), Sustained ventricular arrhythmias. | Based on the information provided, the following differential diagnoses cannot be excluded:  Atrial fibrillation Atrial flutter Atrioventricular nodal reentrant tachycardia (AVNRT) Medication or substance use Nonsustained ventricular tachycardia (NSVT) Premature atrial complexes (PACs) Premature ventricular complexes (PVCs) Sustained ventricular arrhythmias Note that the patient's history and physical examination do not provide sufficient evidence to support or exclude the other differential diagnoses. Further testing and evaluation would be needed to establish a definitive diagnosis. | Atrial fibrillation, Atrial flutter, Atrioventricular nodal reentrant tachycardia (AVNRT), Premature atrial complexes (PACs), Premature ventricular complexes (PVCs), and Nonsustained ventricular tachycardia (NSVT) cannot be excluded based on the history and physical examination in this patient. |
| M14_MANG1 | The following steps are essential to do immediately: ECG, Event monitor, or 24-h Holter monitor. | The following steps are essential to do immediately in this patient:  ECG: An electrocardiogram (ECG) is the most immediate and important test to perform in order to diagnose a cardiac arrhythmia or to rule out an underlying cardiac cause of the patient's symptoms.  Event monitor: An event monitor can be used to capture symptoms in real time, which can help diagnose an arrhythmia that may not be present during an ECG in the doctor's office.  Admit to monitored hospital bed: Based on the patient's symptoms, it may be necessary to admit the patient to a monitored hospital bed for further monitoring and observation. This is especially true if the patient's ECG and event monitor findings are concerning for a serious arrhythmia or if the patient is experiencing symptoms such as light-headedness or syncope.  Note that the other tests, such as a 24-h Holter monitor, cardiac enzymes, and stress test, may also be necessary, but their priority would depend on the results of the ECG and event monitor and the patient's overall clinical condition. | ECG 24-h Holter monitor Admit to monitored hospital bed |
| M14_DX | At this time, the most likely diagnosis cannot be determined based on the information provided. Further evaluation through ECG, Event monitor, or 24-h Holter monitor is necessary to arrive at a definite diagnosis. | Based on the history and physical examination of the patient, the most likely diagnosis at this time is premature atrial complexes (PACs). | Based on the history and physical examination findings, the most likely diagnosis at this time is premature atrial complexes (PACs). |
| M14_MANG2 | Instruct the patient to go to the emergency department right away. | Instruct the patient to go to the emergency department right away. | Instruct the patient to go to the emergency department right away. |
| M14_MANG3 | Immediate synchronized cardioversion and Heparin 60 U/kg IV followed by 12 mg/kg/h drip are essential steps to do immediately. | -Immediate synchronized cardioversion -Aspirin 325 mg daily -Heparin 60 U/kg IV followed by 12 mg/kg/h drip -Transthoracic echocardiography (TTE) -Complete metabolic panel -Serum PT/PTT | Immediate synchronized cardioversion, Diltiazem 15 mg by IV push followed by diltiazem drip at 5 mg/h, and Heparin 60 U/kg IV followed by 12 mg/kg/h drip are essential steps to do immediately. |
| M14_Q1 | Age, history of heart failure, hypertension, peripheral vascular disease, renal function, smoking, type 2 diabetes mellitus, and stroke/transient ischemic attack are used to assess stroke risk in patients with atrial fibrillation. | Age, history of heart failure, hypertension, peripheral vascular disease, smoking, type 2 diabetes mellitus, and stroke/transient ischemic attack are all characteristics used to assess stroke risk in patients with atrial fibrillation. | The characteristics used to assess stroke risk in patients with atrial fibrillation include:  Age History of heart failure Hypertension Smoking Type 2 diabetes mellitus Stroke/transient ischemic attack |
| M14_Q2 | The following are contraindications to the use of direct oral anticoagulant drugs in patients with atrial fibrillation:  Severe mitral stenosis Renal dysfunction with creatinine clearance of 50 mL/min | The following are contraindications to the use of direct oral anticoagulant drugs in patients with atrial fibrillation:  Severe renal dysfunction with creatinine clearance of <30 mL/min Severe liver dysfunction Active bleeding or a high risk of bleeding Significant risk of noncompliance with medication or follow-up. | Severe mitral stenosis and Renal dysfunction with creatinine clearance of 50 mL/min are contraindications to the use of direct oral anticoagulant drugs in patients with atrial fibrillation. Age >65 and Chronic obstructive pulmonary disease (COPD) are not necessarily contraindications, but they may affect the dosing and monitoring of these drugs. |
| M14_Q3 | Intracranial hemorrhage is a contraindication to anticoagulation therapy. | Intracranial hemorrhage is a contraindication to anticoagulation therapy. | Intracranial hemorrhage is a contraindication to anticoagulation therapy. |
| M15_DIFF | The 54-year-old man presents with symptoms of headache, ringing sensation in his right ear, and dizziness. He denies visual symptoms, focal motor or sensory deficits, or nausea and vomiting. He has a 10-year history of migraine headaches and had a head injury in high school but no current symptoms. No concerning findings on physical examination. Further evaluation is necessary to determine the cause of the patient's symptoms, which could be related to a new onset of migraine, Meniere's disease, vestibular neuronitis, or other causes of dizziness. A referral to a neurologist or an ear, nose, and throat specialist may be necessary. | The patient is a 54-year-old man presenting with symptoms of dizziness, headache, and ringing sensation in the right ear. These symptoms have been worsening over the past 3 weeks and are affecting his balance while walking and riding a bicycle. The patient has a history of classical migraine, which was treated with sumatriptan, but this medication did not help with his current symptoms. There is no history of visual disturbances, no nausea or vomiting, and no recent international travel. The patient's review of systems and physical examination are generally noncontributory. The patient is employed, adheres to a healthy lifestyle, and is married with one daughter. Neurological examination shows normal eye movement, normal ear examination, and unsteady gait. A Weber tuning fork test lateralizes to the left ear, and mastoid bone conduction is superior to air conduction on the right side. | Acoustic neuroma, Benign paroxysmal vertigo, Labyrinthitis, Meniere disease, Migraine, Vestibular neuronitis. |
| M15_DIAG | Lumbar puncture and CT scan of the head without contrast are the most appropriate next steps. | Lumbar puncture and CT scan of the head without contrast are the most appropriate next steps. | Auditory testing and MRI of the head with gadolinium contrast are the most appropriate next steps. |
| M15_DX | The most likely diagnosis based on the given results is Acoustic Neuroma, right side. | Based on the provided information, the most likely diagnosis is Acoustic neuroma, right side. | Acoustic neuroma, right side. |
| M15_MANG | Surgical excision of the tumor. | Surgical excision of the tumor is the most appropriate management for a suspected acoustic neuroma, given the presence of a 2-cm mass at the internal auditory meatus. Observation may be appropriate in some cases of small acoustic neuromas with low growth potential, but the size and location of this mass warrant surgical intervention. Radiation therapy is typically not the first-line treatment for acoustic neuromas, but may be considered in cases where surgical excision is not possible or if there are specific medical contraindications to surgery. Aspirin and/or NSAIDs are not typically used for management of acoustic neuromas | Surgical excision of the tumor |
| M15_Q1 | Complications of surgical removal of acoustic neuroma can include:  Facial nerve injury Hearing loss Cerebrospinal fluid (CSF) leak Glossopharyngeal nerve injury Tumor recurrence | Complications of surgical removal of acoustic neuroma may include:  Hearing loss Facial nerve injury Cerebrospinal fluid (CSF) leak Glossopharyngeal nerve injury. | Hearing loss, Facial nerve injury, and Cerebrospinal fluid (CSF) leak are the potential complications of surgical removal of acoustic neuroma. |
| M15_Q2 | The symptoms associated with Meniere disease are hearing loss, tinnitus, vertigo, and sometimes vomiting. | The following are symptoms associated with Meniere disease:  Hearing loss Tinnitus Vertigo Vomiting | Hearing loss, Tinnitus, Vertigo, and Vomiting are symptoms associated with Meniere disease. |
| M15_Q3 | Surgical excision, Stereotactic radiotherapy, and Observation | Chemotherapy, Observation, Stereotactic radiotherapy, and Surgical excision may all be used as treatments for symptomatic acoustic neuromas. | Observation, surgical excision, and stereotactic radiotherapy are treatments that may be used for symptomatic acoustic neuromas. Chemotherapy is not typically used to treat acoustic neuromas. |
| M16_DIFF | At this time, neither Cushing disease nor Diabetic ketoacidosis (DKA) can be excluded as causes of the patient's elevated blood glucose and blood pressure based on the information given in the history and physical examination. Further testing such as plasma glucose and ketones, electrolytes, and beta-hydroxybutyrate levels may be needed to rule out DKA, and cortisol levels and imaging studies may be needed to evaluate for Cushing disease. | Based on the information provided, the following causes cannot be excluded:  Medication non-adherence Metabolic syndrome It's possible the patient's elevated blood glucose and blood pressure are due to her not taking her medications as prescribed or due to having metabolic syndrome, a group of risk factors that increase the risk of developing cardiovascular disease and type 2 diabetes. | Based on the information provided, Cushing disease, diabetic ketoacidosis (DKA), medication non-adherence, metabolic syndrome, pheochromocytoma, sepsis, and urinary tract infection (UTI) cannot be excluded as causes of the patient's elevated blood glucose and blood pressure. |
| M16_DIAG | Basic metabolic panel (BMP) and serum creatinine. | HbA1C and basic metabolic panel (BMP) and serum creatinine. | It is not possible to determine which studies are appropriate without more information about the patient's condition, symptoms, and medical history. A healthcare professional who has access to the patient's complete medical record is best equipped to make this determination. |
| M16_MANG1 | Discuss with the patient the importance of lowering her hemoglobin A1C and blood pressure to avoid long-term complications of kidney failure, heart attack, and stroke. | The following are appropriate next steps:  Discuss with the patient the importance of lowering her hemoglobin A1C and blood pressure to avoid long-term complications of kidney failure, heart attack, and stroke.  Amlodipine 5 mg orally once a day  Losartan 50 mg orally once a day  Empagliflozin 10 mg orally once a day  Glargine insulin 10 U subcutaneously at bedtime | Discuss with the patient the importance of lowering her hemoglobin A1C and blood pressure to avoid long-term complications of kidney failure, heart attack, and stroke.  Empagliflozin 10 mg orally once a day  Losartan 50 mg orally once a day  Lisinopril 80 mg orally once a day  Re-evaluate in 3 months |
| M16_MANG2 | Return to office in 3 months for reevaluation Urine microalbumin screening annually Annual ophthalmologic evaluation Annual podiatric evaluation | The most appropriate discharge instructions include:  Return to office in 3 months for reevaluation Urine microalbumin screening annually Nutritionist evaluation | Return to office in 3 months for reevaluation Urine microalbumin screening annually Podiatric evaluation annually. |
| M16_MANG3 | Fluconazole 150 mg, one dose orally and Metronidazole 500 mg orally for 7 days. | Metronidazole 500 mg orally for 7 days and Fluconazole 150 mg, one dose orally. | Fluconazole 150 mg, one dose orally and Metronidazole 500 mg orally for 7 days. |
| M16_Q1 | Increased urinary excretion of glucose | Increased urinary excretion of glucose. | Increased urinary excretion of glucose. |
| M16_Q2 | Cardiomyopathy | Cardiomyopathy | Cardiomyopathy |
| M17_DIFF | Based on the history and physical examination, the following differential diagnoses cannot be excluded:  Chronic rhinosinusitis Myasthenia gravis Sinus cancer It is important to note that this list is not exhaustive and other diagnoses may also be considered based on further evaluation. | Based on the history and physical examination, the following differential diagnoses cannot be excluded:  Chronic rhinosinusitis Myasthenia gravis It is important to note that the patient has a history of sinusitis and underwent sinus surgery, so chronic rhinosinusitis remains a possibility. The presence of occasional shooting pains in the back of the head and occasional nasal regurgitation of liquids suggest that further evaluation for Myasthenia gravis may be warranted, as these can be symptoms of this condition. Further diagnostic testing, such as laboratory tests and imaging studies, may be necessary to rule out or confirm these diagnoses. | Based on the history and physical examination provided, the following differential diagnoses cannot be excluded:  Chronic rhinosinusitis Adverse effect of inhaled corticosteroids Myasthenia gravis It should be noted that a definitive diagnosis can only be made after further investigation, such as imaging studies, blood work, and possibly nerve conduction studies or a biopsy. |
| M17_DIAG | Given the previous findings, the following steps are most appropriate for guiding diagnosis and treatment:  EMG with repetitive nerve testing Serum acetylcholine receptor (AChR) antibody These tests can help determine if the patient has a neurogenic cause for her symptoms, specifically myasthenia gravis. Other tests, such as MRI of the brain, EEG, muscle biopsy, or serum creatine kinase may be considered if needed based on the results of these initial tests and | Edrophonium test and EMG with repetitive nerve testing. | Based on the information provided, the following steps are most appropriate for guiding diagnosis and treatment:  EMG with repetitive nerve testing Serum acetylcholine receptor (AChR) antibody Edrophonium test These tests can help diagnose conditions such as myasthenia gravis, which is a differential diagnosis that cannot be excluded based on the history and physical examination. Other tests, such as an MRI of the brain, EEG, carotid Doppler ultrasonography, muscle biopsy, and serum creatine kinase, are not directly relevant to the patient's symptoms and may not be necessary for diagnosis at this time. |
| M17_DX | Based on the EMG results with repetitive nerve stimulation and the serum acetylcholine receptor antibody titer being above the normal reference range, the most likely diagnosis is Myasthenia gravis. | Based on the results of the EMG and serum tests, the most likely diagnosis is Myasthenia Gravis. The EMG findings of a decremental response to 3-Hz stimulation and facilitation after 10 sec of exercise are consistent with the findings typically seen in Myasthenia Gravis. The elevated level of acetylcholine receptor antibodies in the serum is also supportive of a diagnosis of Myasthenia Gravis. | The most likely diagnosis based on the results is Myasthenia gravis. The EMG results showing a decremental response to 3-Hz stimulation, less pronounced after 10 sec of exercise (facilitation), and more pronounced after 60 sec of exercise (postfacilitation exhaustion) suggest myasthenic involvement. The elevated level of serum acetylcholine receptor (AChR) antibody (0.28 nmol/L) is also consistent with a diagnosis of Myasthenia gravis. |
| M17_MANG | Based on the findings, the most appropriate steps for guiding diagnosis and treatment would be:  Prednisone 20 mg po once/day Pyridostigmine 60 mg po q 4-6 h Admit the patient for plasma exchange Admit the patient for IV immunoglobulin (IVIG) treatment | Admit the patient for plasma exchange, Prednisone 20 mg po once/ day, Pyridostigmine 60 mg po q 4-6 h. | Based on the results of the EMG with repetitive nerve testing and the serum acetylcholine receptor antibody titer, the most likely diagnosis is Myasthenia Gravis. For this condition, the following steps are most appropriate for guiding diagnosis and treatment:  Admit the patient for plasma exchange Admit the patient for IV immunoglobulin (IVIG) treatment Prednisone 20 mg po once/ day Pyridostigmine 60 mg po q 4-6 h Azathioprine 50 mg po once/ day |
| M17_Q1 | Clindamycin and Botox injections should be avoided in patients with myasthenia gravis. | Clindamycin and Levoquin should be avoided in patients with myasthenia gravis. | Clindamycin and Botox injections should be avoided in patients with myasthenia gravis. |
| M17_Q2 | Graves disease | None of the listed autoimmune diseases can be mistaken for ocular myasthenia. However, the following conditions can mimic ocular myasthenia and should be considered in the differential diagnosis:  Internuclear ophthalmoplegia Infranuclear ophthalmoplegia Thyroid eye disease Myopathic disorders Orbital pseudotumor Migraine. | Graves disease |
| M17_Q3 | IV immunoglobulin, plasma exchange, prednisone, and azathioprine can alter the natural progression of myasthenia gravis. | Plasma exchange, IV immunoglobulin, Azathioprine, Mycophenolate mofetil, Prednisone, and Pyridostigmine can alter the natural progression of myasthenia gravis. | All of the following treatments can alter the natural progression of myasthenia gravis:  Azathioprine IV immunoglobulin Mycophenolate mofetil Plasma exchange Prednisone Pyridostigmine |
| M18_DIFF | Testicular torsion, epididymitis/orchitis, and testicular cancer are the most likely differential diagnoses. | Epididymitis/orchitis, Testicular torsion, Testicular cancer | The most likely differential diagnoses at this time are epididymitis/orchitis and testicular torsion. |
| M18_DIAG | Doppler ultrasonography of the scrotum and urinalysis are the most appropriate immediate steps to order. | Doppler ultrasonography of the scrotum, Surgical exploration of the scrotum, Basic metabolic profile (BMP), Complete blood count (CBC), Urinalysis | The most appropriate immediate steps to order are Doppler ultrasonography of the scrotum and possibly a urinalysis. A surgical exploration of the scrotum may be necessary based on the results of the ultrasound. A basic metabolic profile (BMP) and complete blood count (CBC) may be ordered if deemed necessary. A urethral swab for sexually transmitted infection may be considered if indicated by the patient's history. |
| M18_DX | Testicular torsion. | Based on the results of the urinalysis and Doppler ultrasonography of the scrotum, the most likely diagnosis is Testicular Torsion. The left testis appears heterogeneous and there is no demonstrated venous or arterial Doppler blood flow, which are findings consistent with testicular torsion. | Testicular torsion |
| M18_MANG | Consult a urologist for immediate surgical exploration. | Consult a urologist for immediate surgical exploration | Consult a urologist for immediate surgical exploration |
| M18_Q1 | 12 to 18 yr | 12 to 18 yr | 12 to 18 yr |
| M18_Q2 | Bell-clapper deformity | Bell-clapper deformity is most associated with increased risk for intravaginal testicular torsion. | Bell-clapper deformity and patent processus vaginalis are the deformities most associated with increased risk for intravaginal testicular torsion. |
| M18_Q3 | The average time after onset of pain when testicular salvage rates drop close to zero percent is 6 hours. | Testicular torsion can quickly cause ischemic necrosis of the testis. On average, the testicular salvage rates drop close to zero percent within 6 hours of onset of pain. | 6 h |
| M19_DIFF | Based on the history and physical examination in the given patient, the following differential diagnoses cannot be excluded:  Appendicitis Bowel obstruction Cholecystitis Diverticulitis Pancreatitis Splenic laceration or contusion. | Based on the history and physical examination in this patient, the following differential diagnoses cannot be excluded:  Appendicitis Bowel obstruction Cholecystitis Diverticulitis Pancreatitis Nephrolithiasis Splenic laceration or contusion Urinary tract infection (UTI) | Based on the history and physical examination of the patient, the differential diagnoses that cannot be excluded are:  Appendicitis Bowel obstruction Cholecystitis Diverticulitis Ischemic colitis Nephrolithiasis Pancreatitis Splenic laceration or contusion |
| M19_DIAG | CBC, BMP, urine culture, and urinalysis are the most appropriate steps to guide diagnosis and treatment. | Given the information provided, I'm unable to determine the most appropriate steps to guide diagnosis and treatment. A more detailed history and physical examination is necessary to determine the specific symptoms and signs the patient is experiencing, which can then inform the appropriate diagnostic tests and treatments. | Urinalysis, Urine culture, CBC, and BMP. |
| M19_DX | Based on the results of the urinalysis and CBC, the most likely diagnosis is UTI. | Based on the results of the urinalysis, the most likely diagnosis is UTI (Urinary Tract Infection). The negative nitrite and bacteria results from the urinalysis suggest a UTI. | Based on the results of the urinalysis, CBC, and basic metabolic profile, the most likely diagnosis is UTI (Urinary Tract Infection). The yellow color and low pH of the urine, as well as the presence of 20-25 RBCs/HPF and negative nitrites suggest a possible urinary tract infection. The negative bacteria result does not rule out the possibility of a UTI, as other types of tests may be needed to confirm the presence of an infection. |
| M19_MANG | Based on the results of the urinalysis, CBC, and basic metabolic profile, it appears that the most likely diagnosis is a urinary tract infection (UTI). The most appropriate management steps for UTI would include:  Discharging the patient from the emergency department with a urine strainer Antibiotic treatment, such as ciprofloxacin 500 mg po bid for 10 days Pain management, such as oxycodone/acetaminophen 5 mg/325 mg po q 6 h for 7 days. | Given the results of the Urinalysis and CBC, the most likely diagnosis is a urinary tract infection (UTI). The most appropriate management steps for a UTI include:  Discharging from the emergency department with a urine strainer Ciprofloxacin 500 mg po bid for 10 days Ondansetron 4 mg IV to help with nausea, if needed Oxycodone / acetaminophen 5 mg/ 325 mg po q 6 h for 7 days to manage pain, if needed. | Based on the available information, the most appropriate management steps would be:  Admit the patient to the hospital Ureteroscopic stone removal Shock wave lithotripsy Ciprofloxacin 500 mg po bid for 10 days 0.9 % saline solution, 1 L IV over 1 h Note: The management plan may change based on the patient's condition, underlying health problems and the severity of symptoms. The suggested options are based on the information provided and should be considered in the context of a thorough evaluation and diagnosis by a healthcare professional. |
| M19_Q1 | Based on the urinalysis results and clinical presentation, the most likely composition of the patient's ureteral stone is Calcium oxalate. | Based on the information provided, it is not possible to determine the exact composition of the patient's ureteral stone. Further testing such as CT scan, X-ray, or analysis of the removed stone may be needed to determine its composition. | Based on the information provided, the composition of the patient's ureteral stone cannot be determined from the laboratory results and physical examination findings alone. Further diagnostic imaging, such as abdominal ultrasonography, CT of the abdomen and pelvis, or ureteroscopy, may be needed to identify the type of stone. The most common types of ureteral stones are calcium oxalate, calcium phosphate, and uric acid, but the composition can vary depending on the individual patient's underlying metabolic and dietary factors. |
| M19_Q2 | Uric acid stones can be dissolved with medical management. | Uric acid is the only stone type that can realistically be dissolved and eliminate the need for surgical intervention. | Uric acid is the only stone type that can realistically be dissolved and thus eliminate the need for surgical intervention. |
| M20_DIFF | Based on the history and physical examination, the differential diagnoses that cannot be excluded are: Abscess, Cellulitis, Deep vein thrombosis (DVT). | Based on the history and physical examination, the following differential diagnoses cannot be excluded:  Cellulitis Deep vein thrombosis (DVT) Necrotizing soft-tissue infection Superficial thrombophlebitis | Based on the history and physical examination, the following differential diagnoses cannot be excluded:  Cellulitis Deep vein thrombosis (DVT) |
| M20_DIAG | It is not possible to determine the most appropriate steps without further information about the patient's condition. The tests recommended would depend on the symptoms and other factors specific to the patient. | Without more context or information about the patient's presentation, it is difficult to determine which tests would be the most appropriate. However, based on the differential diagnoses listed in the previous question, some tests that may be considered include:  Complete blood count (CBC) Blood cultures Doppler ultrasonography of leg (for suspected DVT) Basic metabolic panel (Na+, Cl-, K+, HCO₃-, BUN, creatinine, glucose) Note that the final decision regarding which tests to order will depend on the patient's individual situation and needs, as well as the clinician's professional judgement. | It is not possible to determine the most appropriate immediate steps to order without more information about the patient's symptoms and medical history. A clinical diagnosis alone may not be sufficient, and additional testing such as a basic metabolic panel, blood cultures, complete blood count, chest x-ray, Doppler ultrasonography of leg, ECG, leg x-ray, or skin cultures may be needed to make a complete diagnosis. |
| M20_DX | Based on the results of the tests performed, the most likely diagnosis for this patient is Cellulitis. | Based on the results of the tests and the patient's symptoms, it appears that the most likely diagnosis is cellulitis. | It is not possible to determine the most appropriate steps to order or the most likely diagnosis without more information about the patient's symptoms and medical history. A complete physical examination and relevant diagnostic tests may be necessary to make an accurate diagnosis. |
| M20_MANG | Admit the patient to the hospital, Blood cultures, Cephalexin 500 mg PO every q 6 h, Elevate the right leg while in bed, and Tobramycin 100 mg IV every q 8 h plus piperacillin 3 g IV every q 4 h. | The most appropriate management of this patient's condition would be:  Admit the patient to the hospital Blood cultures Cephalexin 500 mg po every q 6 h Elevate the right leg while in bed Encourage ambulation Ibuprofen 600 mg po every q 6 h as needed for pain | Admit the patient to the hospital, Elevate the right leg while in bed, Blood cultures, Nafcillin 1 g IV every q 6 h, Tobramycin 100 mg IV every q 8 h plus piperacillin 3 g IV every q 4 hr, Vancomycin 1 g IV every q 12 h, and Morphine 8 mg IV every q 6 h as needed for pain. |
| M20_Q1 | Cephalexin and dicloxacillin are appropriate treatments for mild cellulitis. | Cephalexin and dicloxacillin are appropriate treatments for mild cellulitis. | Cephalexin, dicloxacillin, and levofloxacin are appropriate treatments for mild cellulitis. |
| M20_Q2 | The following factors affect the choice of antimicrobial therapy in patients with cellulitis:  High prevalence of MRSA in the community History of hives after taking penicillin Presence of regional adenopathy Size of the area of erythema | The following factors affect the choice of antimicrobial therapy in patients with cellulitis:  High prevalence of MRSA in the community History of hives after taking penicillin Presence of regional adenopathy Size of the area of erythema | High prevalence of MRSA in the community, history of hives after taking penicillin, presence of regional adenopathy, and size of the area of erythema all affect the choice of antimicrobial therapy in patients with cellulitis. |
| M20_Q3 | The factors that suggest that a patient with cellulitis needs to be admitted to the hospital are:  Hypotension Delirium Lymphangitis Low-grade fever with a rapid heart rate or difficulty breathing Large, spreading area of erythema Systemic symptoms such as vomiting and significant pain. | The following factors suggest that a patient with cellulitis needs to be admitted to the hospital:  Hypotension Low-grade fever Lymphangitis Delirium (if severe or if associated with altered mental status or confusion) Leukocytosis Vomiting (if severe or if associated with dehydration) | The following factors suggest that a patient with cellulitis needs to be admitted to the hospital:  Hypotension Delirium Vomiting High fever Rapid progression of the infected area Evidence of systemic toxicity, such as a high white blood cell count (leukocytosis) Presence of co-morbidities that increase the risk of complications, such as immunosuppression or underlying vascular disease. |
| M20_Q4 | The features that help differentiate cellulitis and DVT of the leg are:  Leg swelling Peau d'orange skin (orange peel-like appearance of the skin due to edema) Positive Homan sign (pain and swelling in the calf upon dorsiflexion of the foot) Lymphangitis (red streaks extending from the infected site towards the axilla) High fever Low-grade fever may be present in both cellulitis and DVT, but is more commonly seen in cellulitis. | Leg swelling, peau d'orange skin, positive Homan sign, and lymphangitis are features that can help differentiate cellulitis and DVT of the leg. | Leg swelling, positive Homan sign, and peau d'orange skin are features that suggest a diagnosis of DVT rather than cellulitis. Lymphangitis is a feature that may be present in both cellulitis and DVT. |
| M21_Q1 | Based on the information available in the history and physical examination, the peripheral nerves and spinal cord are most likely to be involved in the patient's symptoms. The symptoms described (such as "novocaine wearing off" feeling in both legs and "funny" feeling in fingertips) are consistent with peripheral neuropathy or spinal cord involvement. The normal cranial nerves, motor exam, and sensation exam support that the cerebral cortical sensory areas and midbrain are unlikely to be involved at this time. The autonomic ganglia may also be involved in some cases of peripheral neuropathy, but further evaluation is needed to determine if this is present in this patient | Based on the history and physical examination, the peripheral nerves and spinal cord are most likely to be involved in this patient's symptoms. | Based on the given history and physical examination, the most likely components of the nervous system that are involved in this patient's symptoms are the peripheral nerves and spinal cord. The patient reports bilateral numbness in her waist to toes which suggest a peripheral nerve or spinal cord issue. The examination shows normal results for cranial nerves, motor and sensory functions, reflexes, coordination, gait, balance, and mental status, which supports the involvement of peripheral nerves and spinal cord in the patient's symptoms. |
| M21_DIFF | Based on the information available in the history and physical examination, several differential diagnoses cannot be excluded at this time, including:  Peripheral neuropathy due to toxins Acute viral infection Multiple sclerosis (MS) Connective tissue disorder Myelopathy due to vitamin deficiency While the normal bladder and bowel function and absence of neck or back pain suggest that lumbosacral plexitis, spinal cord compression by tumor, spinal cord hemorrhage, and spinal epidural abscess are less likely, further evaluation is needed to definitively rule these out. The absence of other symptoms such as rash or joint pain, and a normal review of systems, suggest that sarcoidosis is less likely as well. | Based on the history and physical examination, the following differential diagnoses cannot be excluded: Acute viral infection, peripheral neuropathy due to toxins, multiple sclerosis (MS), spinal cord compression by tumor, spinal cord hemorrhage, and spinal epidural abscess. | Based on the given history and physical examination, the following differential diagnoses cannot be excluded:  Acute viral infection Peripheral neuropathy due to toxins Lumbosacral plexitis Spinal cord compression by tumor Multiple sclerosis (MS) The patient reports bilateral numbness extending from her waist to toes, which could be due to an acute viral infection or a peripheral nerve issue such as peripheral neuropathy due to toxins. The physical exam shows normal results for cranial nerves, motor and sensory functions, reflexes, coordination, gait, and balance, which suggests the involvement of the peripheral nerves and spinal cord in the patient's symptoms. Lumbosacral plexitis is a peripheral nerve issue in the lower back. Multiple sclerosis is a chronic autoimmune disease that can cause symptoms similar to those reported by the patient. The possibility of spinal cord compression by a tumor or spinal cord hemorrhage cannot be ruled out based on the information available. |
| M21_DIAG | MRI of the cervical and thoracic spine, and EMG of the legs. | The most appropriate immediate next steps are MRI of the cervical spine and MRI of the thoracic spine. An MRI can help to assess for spinal cord compression by tumor, spinal cord hemorrhage, or other structural abnormality that could be causing the patient's symptoms. | Based on the information provided, the most appropriate immediate next steps would be:  MRI of the cervical and thoracic spine Electromyography (EMG) of the legs |
| M21_MANG | Given the MRI findings, the most appropriate steps to guide diagnosis and treatment are:  Nerve conduction studies and electromyography Spinal tap | Given the MRI findings, the most appropriate steps to guide diagnosis and treatment would be:  Nerve conduction studies and electromyography Spinal tap CT of the spine (if the MRI didn't give clear picture of spinal cord, to rule out spinal cord compression) EEG and Doppler studies of the legs might be considered based on the specific results of the above mentioned tests and to rule out other potential causes. MRI of the brain would be considered if there is any concern for central nervous system involvement. | Based on the MRI findings, the most appropriate next steps to guide diagnosis and treatment would be:  Nerve conduction studies and electromyography Spinal tap MRI of the brain |
| M21_DX | Multiple sclerosis is the most likely diagnosis based on the MRI findings mentioned. | Based on the information available, it is not possible to determine the most likely diagnosis without further information and tests. | I'm sorry, I cannot determine the most likely diagnosis based on the information provided. It would require a full evaluation and interpretation of the patient's clinical history, physical examination, and imaging findings, along with laboratory and other diagnostic tests. A healthcare professional, such as a neurologist, would be best equipped to make a definitive diagnosis. |
| M21_MANG2 | The most appropriate initial therapies for multiple sclerosis (MS) can include:  Interferon beta-1a, 30 mcg IM once/wk Dimethyl fumarate 120 mg po bid for 7 days Natalizumab 300 mg IV q 4 wk | Azathioprine, Dimethyl fumarate, Interferon beta-1a, Methylprednisolone, Mitoxantrone, and Natalizumab can all be considered initial therapies for multiple sclerosis. | The following can be considered initial therapies for this patient:  Interferon beta-1a, 30 mcg IM once/wk Dimethyl fumarate 120 mg po bid for 7 days Natalizumab 300 mg IV q 4 wk Methylprednisolone 1 g IV once per day for 3 d |
| M21_Q1 | Transverse myelitis is a neurological condition characterized by symptoms and signs including:  Weakness below the level of the lesion Sensory loss below the level of the lesion Sphincteric and sexual dysfunction | The symptoms and signs of transverse myelitis include:  Weakness below the level of the lesion Sensory loss below the level of the lesion Sphincteric and sexual dysfunction Visual loss Dysarthria (slurred speech) Dysphasia (difficulty speaking) | Transverse myelitis is a neurological disorder characterized by inflammation of the spinal cord, which can result in the following symptoms and signs:  Weakness below the level of the lesion Sensory loss below the level of the lesion Sphincteric and sexual dysfunction Visual loss Dysarthria (difficulty speaking) |
| M21_Q2 | Living in a temperate climate during the first 15 years of life Female sex HLA-DR2 allotype Cigarette smoking | Living in a temperate climate during the first 15 years of life Female sex Human leukocyte antigen (HLA) DR2 allotype Cigarette smoking | The following factors are associated with a higher risk of developing multiple sclerosis:  Female sex Living in a temperate climate during the first 15 years of life Human leukocyte antigen (HLA) DR2 allotype Cigarette smoking |
| M21_Q3 | Optic neuritis and internuclear ophthalmoplegia are commonly seen in patients with multiple sclerosis (MS). | Optic neuritis, and internuclear ophthalmoplegia are commonly seen in patients with multiple sclerosis (MS). | Optic neuritis and Internuclear ophthalmoplegia are commonly seen in patients with multiple sclerosis (MS). |
| M21_Q4 | The most common subtype of multiple sclerosis is Relapsing - remitting. | The most common subtype of multiple sclerosis is relapsing-remitting | The most common subtype of multiple sclerosis is relapsing-remitting multiple sclerosis. |
| M22_DIFF | Opioid overdose, Tricyclic antidepressant (TCA) overdose, Calcium channel blocker overdose, Beta-blocker overdose, Digoxin overdose. | Opioid overdose and TCA overdose. | Opioid overdose, Beta-blocker overdose, Calcium channel blocker overdose, Tricyclic antidepressant (TCA) overdose. |
| M22_MANG1 | Cardiac monitoring  Naloxone 0.4 mg IV and monitoring for improvement in symptoms.  Bedside glucose level  Chest x-ray | Cardiac monitoring  Naloxone 0.4 mg IV and monitoring for improvement in symptoms.  Bedside glucose level  Serum electrolyte concentrations | Naloxone 0.4 mg IV and monitoring for improvement in symptoms.  Cardiac monitoring  Bedside glucose level  Blood cultures  Chest x-ray |
| M22_DIFF | Opioid overdose. | Opioid overdose | Opioid overdose. |
| M22_MANG2 | Admit the patient to the general medicine floor, Cardiac pacing, Atropine 0.5 to 1 mg IV q 3 to 5 min, Epinephrine 1 mcg/min IV, Glucagon 5 mg over 1 min IV followed by 2 to 5 mg/h IV infusion. | Cardiac pacing, Atropine 0.5 to 1 mg IV q 3 to 5 min, Epinephrine 1 mcg/min IV, and Glucagon 5 mg over 1 min IV followed by 2 to 5 mg/h IV infusion. | Admit the patient to the general medicine floor, Atropine 0.5 to 1 mg IV q 3 to 5 min, and Epinephrine 1 mcg/min IV |
| M22_Q1 | Bradycardia, Dilated pupils, Lethargy, Hypotension, Tachypnea. | Opioid overdose typically presents with:  Bradycardia Dilated pupils Lethargy Hypotension Respiratory depression or shallow breathing (bradypnea or tachypnea) | Bradycardia, dilated pupils, lethargy, hypotension, and respiratory depression (tachypnea or bradypnea) are typically present in opioid overdose. |
| M22_Q2 | The following can be used in treatment of beta-blocker overdose:  -High-dose insulin (bolus of 1 unit/ kg of regular, short-acting insulin, then 0.5 units/ kg / h IV) and glucose therapy titrated to maintain normal glucose levels -Lipid emulsion therapy 1.5 mL/ kg of 20% solution over 2 min, then 1.5 mL/ kg over 60 min | High-dose insulin (bolus of 1 unit/kg of regular, short-acting insulin, then 0.5 units/kg/h IV) and glucose therapy titrated to maintain normal glucose levels, Lipid emulsion therapy 1.5 mL/kg of 20% solution over 2 min, then 1.5 mL/kg over 60 min | High-dose insulin (bolus of 1 unit/ kg of regular, short - acting insulin, then 0.5 units/ kg / h IV) and glucose therapy titrated to maintain normal glucose levels  Calcium gluconate 1 g via peripheral IV  Lipid emulsion therapy 1.5 mL/ kg of 20% solution over 2 min, then 1.5 mL/ kg over 60 min |
| M22_Q3 | The typical symptoms and signs of the Cushing reflex are:  Hypertension Bradycardia Widened pulse pressure Respiratory depression | Hypertension, bradycardia, and respiratory depression are typical symptoms and signs of the Cushing reflex. | Hypertension Bradycardia Widened pulse pressure Respiratory depression |
| M23_DIFF | Based on the history and physical examination, the following differential diagnoses cannot be excluded: Intracerebral hemorrhage, Stroke affecting the posterior circulation, Seizure, Cardiac arrhythmia, Orthostatic hypotension, Subarachnoid hemorrhage, Vasovagal syncope, Opioid overdose, Hypoglycemia, Alcohol intoxication/withdrawal. | Based on the information provided in the history and physical examination, the following differential diagnoses cannot be excluded at this time:  Stroke affecting the posterior circulation Intracerebral hemorrhage Subarachnoid hemorrhage Seizure Cardiac arrhythmia Alcohol withdrawal Opioid overdose Meningitis It is important to note that this list is not exhaustive and other diagnoses may need to be considered based on further examination and test results. | Based on the history and physical examination, the following differential diagnoses cannot be excluded:  Stroke affecting the posterior circulation Intracerebral hemorrhage Subarachnoid hemorrhage Seizure Cardiac arrhythmia Opioid overdose Orthostatic hypotension Pulmonary embolism (PE) |
| M23_MANG1 | The following steps are essential to do immediately: Cardiac monitoring, BP measurement every 5 min until stable, Pulse oximetry, Dextrose 50 g IV (if hypoglycemia is suspected), Naloxone 0.4 mg IV (if opioid overdose is suspected), Bedside glucose measurement (if hypoglycemia is suspected), Toxicology screening. | Based on the information provided, the following steps are essential to do immediately:  BP measurement every 5 minutes until stable Pulse oximetry Cardiac monitoring Toxicology screening Naloxone 0.4 mg IV (if opioid overdose is suspected) The patient's vital signs, including blood pressure and heart rate, need to be closely monitored and stabilized as soon as possible. Pulse oximetry should be used to monitor the patient's oxygen saturation levels. If there is suspicion of opioid overdose, administration of Naloxone is indicated. A toxicology screening should also be performed to determine if any other substances may have contributed to the patient's symptoms. These interventions are necessary to stabilize the patient's condition and guide further diagnostic and therapeutic measures. | Based on the information given, the following steps are essential to do immediately:  BP measurement every 5 minutes until stable Cardiac monitoring Pulse oximetry Naloxone 0.4 mg IV (in case of suspected opioid overdose) Bedside glucose measurement Toxicology screening |
| M23_MANG2 | Given the previous findings, the most appropriate steps to guide diagnosis and treatment are: Reexamination of the patient, Seek additional history from patient's family and records, Head CT, ECG, Serum chemistry profile, Tilt table testing (if orthostatic hypotension is suspected), Carotid Doppler testing (if stroke is suspected), Lumbar puncture for examination of CSF (if subarachnoid hemorrhage is suspected), CBC. | Based on the previous findings, the following steps would be appropriate to guide diagnosis and treatment:  Head CT: Given the patient's history of sudden onset of unconsciousness and confusion, a head CT would be appropriate to rule out intracerebral or subarachnoid hemorrhage, which could present with similar symptoms.  Reexamination of the patient: The patient's condition has changed since initial examination, with extreme lethargy and a decrease in vital signs. It is important to regularly reassess the patient's condition to guide appropriate management.  Seek additional history from patient's family and records: As the patient is unable to provide a comprehensive medical history, it is important to seek additional information from the patient's family or medical records to identify any potential underlying medical conditions or risk factors that could contribute to the patient's symptoms.  Serum chemistry profile: A serum chemistry profile can help assess for metabolic disturbances that could be contributing to the patient's symptoms.  ECG: Although a normal sinus rhythm was noted on the patient's initial ECG, it is important to regularly monitor the patient's heart rhythm and to repeat the ECG as necessary.  These steps would provide more information to help guide a definitive diagnosis and appropriate treatment for the patient. | Based on the normal results from cardiac monitoring, pulse oximetry, ECG, and bedside glucose, the following steps are the most appropriate to guide diagnosis and treatment:  Reexamination of the patient Seek additional history from the patient's family and records Head CT Lumbar puncture for examination of CSF (considering the findings from the initial examination) Carotid Doppler testing (considering the possibility of a stroke) Serum chemistry profile CBC ECG Tilt table testing (considering the possibility of orthostatic hypotension) Toxicology screen (as part of a comprehensive evaluation) |
| M23_DX1 | Alcohol withdrawal Seizure Subarachnoid hemorrhage Vasovagal syncope | Based on the new information, the likely differential diagnoses now include:  Alcohol withdrawal Seizure Vasovagal syncope Intracerebral hemorrhage Subarachnoid hemorrhage Note: Although the patient's medical history is noncontributory and the cardiac monitor has shown normal sinus rhythm, cardiac arrhythmia cannot be excluded and should be considered in the differential diagnoses. | Alcohol withdrawal Seizure Subarachnoid hemorrhage |
| M23_DIAG | Repeat physical examination of patient for signs of bleeding Plasma fibrinogen and d-dimer levels Liver function tests HIV testing Hepatitis C testing | Repeat physical examination of patient for signs of bleeding | Repeat physical examination of patient for signs of bleeding |
| M23_DX2 | Given the findings of thrombocytopenia and petechiae on the lower legs and an area of ecchymosis on the upper back, the most likely diagnosis at this time is Immune thrombocytopenia (ITP). | Based on the results of additional testing, the most likely diagnosis is Immune thrombocytopenia (ITP). | The most likely diagnosis is Immune thrombocytopenia (ITP). |
| M23_MANG3 | Admit the patient to the hospital Platelet transfusion 6 U IV Prednisone 1 mg/kg po Consider referral to a hematologist for further evaluation and management. | Admit the patient to the hospital, Platelet transfusion 6 U IV, Prednisone 1 mg/kg po, Immune globulin 1 g/kg IV. | The most appropriate management options at this time are:  Admit the patient to the hospital Platelet transfusion 6 U IV Immune globulin 1 g/kg IV Prednisone 1 mg/kg po The patient has thrombocytopenia, and admission to the hospital is necessary to monitor the patient and ensure proper management. Platelet transfusion and immune globulin are treatments for thrombocytopenia, while prednisone can help manage the underlying condition causing the thrombocytopenia. |
| M23_Q1 | Onset of syncope immediately after standing up and Multiple recurrences of syncope within a short time can suggest a dangerous cause of syncope. | All of the options listed could suggest a dangerous cause of syncope:  Associated heart murmur: could indicate an underlying cardiac issue Onset of syncope during exertion: could indicate cardiac ischemia or arrhythmia Onset of syncope during strong emotional display: could indicate a cardio-inhibitory response Onset of syncope immediately after standing up: could indicate a neuro-cardiogenic response Multiple recurrences of syncope within a short time: could indicate an ongoing cardiac or neurological issue Syncope preceded by diaphoresis: could indicate a neuro-cardiogenic response | The following features would suggest a dangerous cause of syncope:  Onset of syncope during exertion Associated heart murmur Multiple recurrences of syncope within a short time Syncope preceded by diaphoresis |
| M23_Q2 | Warfarin, heparin, and trimethoprim/sulfamethoxazole are more often associated with the development of thrombocytopenia. | Heparin, trimethoprim / sulfamethoxazole, and quinine are more often associated with the development of thrombocytopenia. | Heparin, quinine, and trimethoprim/sulfamethoxazole are more often associated with the development of thrombocytopenia. |
| M23_Q3 | The following infections are associated with development of thrombocytopenia:  Epstein-Barr virus infection HIV infection Lyme disease Syphilis Hepatitis C Trichinosis | The following infections are associated with the development of thrombocytopenia:  Epstein-Barr virus infection HIV infection Lyme disease Hepatitis C | The following infections are associated with development of thrombocytopenia:  Epstein-Barr virus infection HIV infection Lyme disease Syphilis Trichinosis |
| M23_Q4 | A subarachnoid hemorrhage is the least likely type of stroke to cause loss of consciousness (LOC). | Anterior circulation ischemic stroke is the least likely type of stroke to cause loss of consciousness (LOC). | Anterior circulation ischemic stroke is the least likely to cause loss of consciousness (LOC). |
| M24_DIFF | Based on the history and physical examination, the following diagnoses cannot be excluded:  Psychogenic polydipsia Diabetes insipidus Hypercalcemia. | DIFF: Diabetes Mellitus, Diabetes Insipidus, Hypercalcemia, Psychogenic Polydipsia, Urinary Tract Infection, Drug Ingestion | Based on the history and physical examination, the following diagnoses cannot be excluded:  Hypercalcemia Diabetes insipidus Psychogenic polydipsia |
| M24_DIAG1 | Based on the symptoms, the following studies would be most appropriate:  Serum electrolyte concentrations Serum calcium and phosphate Serum liver function studies (LFT) Serum TSH and free T4 Serum urea and creatinine Serum and urine osmolality Random blood glucose Complete blood count (CBC) Serum C-reactive protein (CRP) A CT scan of the chest, abdomen, and pelvis would not be necessary at this time based on the limited information given. | DIAG1: Random blood glucose, Serum electrolyte concentrations, Serum calcium and phosphate, Serum TSH and free T4, Serum urea and creatinine, Serum and urine osmolality. | The most appropriate studies to obtain at this time based on the history and physical examination are:  Serum electrolyte concentrations Serum calcium and phosphate Serum urea and creatinine Serum and urine osmolality These tests can help to diagnose or rule out conditions such as hypercalcemia, diabetes insipidus, or psychogenic polydipsia, which are possible based on the history and physical examination. |
| M24_DX | Based on the results provided, differential diagnoses that best account for the patient's findings include:  Nephrogenic diabetes insipidus Psychogenic polydipsia | Based on the results, none of the conditions seem to account for the patient's findings. The serum glucose levels (112 mg/dL) and creatinine (0.8 mg/dL) levels are normal. The TSH (0.45 mU/L) and FT4 (1.2 ng/dL) levels suggest normal thyroid function. The serum calcium levels (13.6 mg/dL) are normal and do not indicate hypercalcemia. The serum electrolyte levels (Na+, K+, Cl-, and HCO3-) and osmolality levels (285 mOsmol/kg) are also normal. The urine osmolality (210 mOsmol/kg) is within the normal range. None of these results support a diagnosis of central diabetes insipidus, diabetes mellitus, hypercalcemia, nephrogenic diabetes insipidus, or psychogenic polydipsia. Further testing may be needed to arrive at a diagnosis. | Based on the results of the testing, the differential diagnoses that best account for the patient's findings are:  Central diabetes insipidus (low urine osmolality with low serum osmolality) Nephrogenic diabetes insipidus (low urine osmolality with normal serum osmolality) |
| M24_Q1 | Addison disease, Excessive calcium consumption, Vitamin D toxicity, Primary hyperparathyroidism, Sarcoidosis, Paraneoplastic syndrome with cancer secreting PTH-related peptide should be evaluated as possible causes of the patient's hypercalcemia. | Primary hyperparathyroidism, Vitamin D toxicity, and bone resorption due to malignant infiltration should be evaluated as possible causes of the patient's hypercalcemia. | Bone resorption due to malignant infiltration, Excessive calcium consumption, Vitamin D toxicity, Primary hyperparathyroidism, Paraneoplastic syndrome with cancer secreting PTH-related peptide. |
| M24_DIAG2 | The most appropriate studies to aid in establishing the diagnosis of hypercalcemia are:  Serum total vitamin D level 24-hour urine calcium Urine calcium excretion ratio Serum parathyroid hormone (PTH) level Ultrasonography of the neck Sestamibi scintigraphy scan of the neck | Serum total vitamin D level Serum parathyroid hormone (PTH) level 24-hour urine calcium Water deprivation test Urine calcium excretion ratio Sestamibi scintigraphy scan of the neck Chest x-ray | Serum calcium, serum PTH, 24-hour urine calcium, Urine calcium excretion ratio, Water deprivation test, Ultrasonography of the neck, Sestamibi scintigraphy scan of the neck. |
| M24_Q2 | Based on the results of the tests, the most likely cause of hypercalcemia in this patient is primary hyperparathyroidism due to a right inferior parathyroid adenoma or cancer. The high level of parathyroid hormone (PTH) and the nodule seen on ultrasonography of the neck suggest the presence of a parathyroid adenoma or cancer. The elevated 24-hour urine calcium also supports this diagnosis. The normal result of chest x-ray and the normal result of protein electrophoresis rules out lymphoma and multiple myeloma, respectively. The normal result of protein electrophoresis also rules out ectopic parathyroid adenoma. The normal level of total vitamin D and the normal result of angiotensin converting enzyme (ACE) suggest that vitamin D toxicity and sarcoidosis are not the cause of hypercalcemia. The normal result of chest x-ray rules out small-cell lung cancer secreting PTH-related peptide. | The most likely cause of hypercalcemia in this patient is Primary hyperparathyroidism due to right inferior parathyroid adenoma or cancer based on elevated parathyroid hormone (PTH) levels and a nodule in the right lower pole of the thyroid on ultrasonography. However, further evaluation, including a biopsy or surgery of the nodule, is required to confirm the diagnosis. | Primary hyperparathyroidism due to right inferior parathyroid adenoma or cancer. |
| M24_MANG | The most appropriate management steps for this patient are:  Right inferior parathyroidectomy Watch and wait, with regular monitoring of serum calcium levels and repeat neck imaging in 6 months Note: Other management options such as Calcitonin and Pamidronate can be used to temporarily control hypercalcemia, but they don't treat the underlying cause of primary hyperparathyroidism and long-term monitoring is still required. The rest of the options mentioned are not directly relevant to the management of primary hyperparathyroidism. | The most appropriate management steps for a patient with hypercalcemia due to primary hyperparathyroidism likely due to right inferior parathyroid adenoma are:  Right inferior parathyroidectomy 3-and-a-half glands parathyroidectomy Watch and wait, with regular monitoring of serum calcium levels and repeat neck imaging in 6 months Calcitonin 5 IU/kg subcutaneously twice a day Pamidronate 60 to 90 mg IV infusion. | The most appropriate management steps for this patient are:  Right inferior parathyroidectomy 3-and-a-half glands parathyroidectomy Watch and wait, with regular monitoring of serum calcium levels and repeat neck imaging in 6 months |
| M24_Q3 | Abdominal pain, confusion, constipation, depression, diarrhea, lethargy, nausea and vomiting, polydipsia and polyuria, reduced appetite, and tetany. | Polydipsia and polyuria, nausea and vomiting, constipation, depression, lethargy, reduced appetite, confusion, and tetany are symptoms of hypercalcemia. | Polydipsia and polyuria, constipation, nausea and vomiting, lethargy, confusion, depression, reduced appetite, abdominal pain, and tetany are symptoms of hypercalcemia. |
| M24_Q4 | Hypercalcemia can be caused by:  Vitamin D intoxication Milk-alkali syndrome Metastatic breast cancer Metastatic prostate cancer Multiple myeloma Lymphoma Neuroendocrine tumor secreting PTH related peptide Parathyroid gland overactivity (Primary hyperparathyroidism) Vitamin D-resistant rickets Thiazide diuretics Granulomatous diseases like sarcoidosis Tumor-induced osteomalacia | Hypercalcemia can be caused by:  Vitamin D intoxication Milk-alkali syndrome Hyperparathyroidism Metastatic breast cancer and metastatic prostate cancer Multiple myeloma Lymphoma Neuroendocrine tumor secreting PTH related peptide Addison disease Tuberculosis Vitamin D intoxication Cushing syndrome Proton pump inhibitors. | Some of the causes of hypercalcemia include:  Vitamin D intoxication Milk-alkali syndrome Hyperparathyroidism (primary or secondary) Malignancy (such as multiple myeloma, metastatic breast or prostate cancer, lymphoma, small-cell lung carcinoma, neuroendocrine tumor secreting PTH related peptide) Sarcoidosis Tuberculosis Proton pump inhibitors. |
| M24_Q5 | Hypocalcemia, recurrent laryngeal nerve palsy, and tetany are possible complications of parathyroidectomy. | Hypocalcemia, recurrent laryngeal nerve palsy, and tetany are possible complications of parathyroidectomy. | Hypocalcemia, recurrent laryngeal nerve palsy, and tetany are possible complications of parathyroidectomy. |
| M25_DIFF | Based on the information provided in the history and physical examination, the following differential diagnoses cannot be excluded:  Breast cancer Fibrocystic breast changes Pregnancy Pituitary tumor Polycystic ovarian syndrome (PCOS) | Based on the information provided, the following differential diagnoses cannot be excluded: -Breast cancer -Fibrocystic breast changes -Polycystic ovarian syndrome (PCOS) -Pregnancy -Pituitary tumor | Breast cancer, fibrocystic breast changes, pituitary tumor, pregnancy, and hypothalamic tumor are common and/or dangerous differential diagnoses that cannot be excluded based on history and physical examination in this patient. |
| M25_DIAG1 | The most appropriate initial steps to guide diagnosis and treatment would be:  Serum β-hCG concentration Urine pregnancy test Serum prolactin (PRL) level Mammography Pelvic ultrasound. | Serum β-hCG concentration, urine pregnancy test, mammography, and pelvic ultrasound. | Urine pregnancy test, Serum β-hCG concentration, Serum prolactin (PRL) level, and Mammography are the most appropriate initial steps to guide diagnosis and treatment. |
| M25_DIAG2 | Given the elevated prolactin level and slightly enlarged sellar mass, the following steps are now most appropriate for guiding diagnosis and treatment:  Humphrey's visual field test Serum ACTH stimulation test Serum cortisol level Serum estradiol level (for females) Serum insulin-like growth factor 1 (IGF-1) level | Bilateral ocular tonometry, Humphrey's visual field test, Serum ACTH stimulation test, Serum free cortisol level. | Based on the results of the previous tests and the suspicion for a pituitary tumor, the following tests would be most appropriate for guiding diagnosis and treatment:  Serum ACTH stimulation test Serum cortisol level (AM cortisol) Bilateral ocular tonometry Humphrey's visual field test |
| M25_DX | Based on the test results, the most likely diagnosis in this patient is hyperprolactinemia due to a pituitary microadenoma. | The most likely diagnosis in this patient is hyperprolactinemia due to a pituitary microadenoma or a pituitary macroadenoma. | Based on the normal results from the Humphrey's visual field test and the normal levels of LH, FSH, free T4, estradiol, IGF-1, and AM cortisol, the most likely diagnosis in this patient is hyperprolactinemia due to a pituitary microadenoma or a pituitary stalk compression by a nonfunctioning pituitary macroadenoma. |
| M25_MANG | Bromocriptine 2.5 mg orally twice a day, Cabergoline 1 mg orally 2 times per week, and Pegvisomant 10 mg subcutaneously daily are appropriate treatments for hyperprolactinemia due to pituitary macroadenoma. | The appropriate treatment for a patient with hyperprolactinemia due to pituitary macroadenoma is:  Bromocriptine 2.5 mg orally twice a day Cabergoline 1 mg orally 2 times per week Pegvisomant 10 mg subcutaneously daily Outpatient pituitary radiotherapy Admit to the hospital for immediate transphenoidal surgical resection of pituitary tumor (if other medical treatments are not effective) | Bromocriptine 2.5 mg orally twice a day Cabergoline 1 mg orally 2 times per week Pegvisomant 10 mg subcutaneously daily |
| M25_Q1 | The most common complications of macroprolactinoma are:  Infertility Bitemporal hemianopia Hirsutism Hypocalcemia | Bitemporal hemianopia, infertility, hypocalcemia, and optic neuropathy are the most common complications of macroprolactinoma. | The most common complications of macroprolactinoma are:  Infertility Bitemporal hemianopia Optic neuropathy |
| M25_Q2 | Prolactinoma is not commonly associated with Carney complex, Cowden syndrome, Familiar hypocalcuric hypercalcemia, or von Hippel-Lindau syndrome. It is most commonly associated with McCune-Albright syndrome and Multiple Endocrine Neoplasia Type 1 (MEN1). | POEMS syndrome, Carney complex, and MEN1 are conditions most commonly associated with prolactinoma. | Prolactinomas are not commonly associated with Carney complex, Cowden syndrome, Familiar hypocalcuric hypercalcemia, McCune-Albright syndrome, POEMS syndrome, and von Hippel-Lindau syndrome. Prolactinomas are more commonly associated with Multiple endocrine neoplasia Type 1 (MEN1). |
| M25_Q3 | The drugs that can cause hyperprolactinemia are:  Metoclopramide Haloperidol Risperidone Verapamil | The drugs that can cause hyperprolactinemia are:  Metoclopramide Haloperidol Risperidone Note: Other medications such as tricyclic antidepressants, some anti-anxiety medications, and certain anti-psychotics can also cause elevated prolactin levels. However, it's important to note that sometimes the cause of hyperprolactinemia may not be related to any medication and could be due to a medical condition such as a pituitary tumor. | Hyperprolactinemia can be caused by the following drugs:  Metoclopramide Haloperidol Risperidone |
| M26_DIFF | Benign prostatic enlargement (BPH) Prostate cancer Urinary tract infection | Based on the patient's symptoms, the following differential diagnoses should be considered:  Benign prostatic enlargement (also called benign prostatic hypertrophy [BPH]) Prostate cancer Urinary tract infection Drug side effects (related to Avanafil) Bladder stone Urethral stricture | Benign prostatic enlargement (BPH), Prostate cancer, Urinary tract infection. |
| M26_DIAG | Post-void residual measurement Serum PSA level Urinalysis and urine culture | The most appropriate next steps at this time would be:  Urinalysis and urine culture Serum PSA level Prostate ultrasound Post-void residual measurement (to assess for bladder outlet obstruction) | Post-void residual measurement, Serum PSA level, Urinalysis and urine culture. |
| M26_DX | Benign prostatic hyperplasia (BPH) | Based on the history and physical examination results, the most likely diagnosis is Benign prostatic hyperplasia (BPH). The patient's symptoms of increased frequency of urination, difficulty starting a stream, and prostate gland on exam without tenderness are consistent with BPH. The serum PSA level of 3.2 ng/mL is within normal limits for a patient of his age. | Benign prostatic hyperplasia (BPH). |
| M26_MANG | Doxazosin 1 mg po daily, Finasteride 5 mg po daily, Oxybutinin 5 mg po bid, and Tadalafil 2.5 mg po daily. | The most appropriate management options for a patient with BPH include:  Doxazosin 1 mg po daily Finasteride 5 mg po daily Oxybutinin 5 mg po bid Transurethral resection of the prostate | Oxybutinin 5 mg po bid and/or Tadalafil 2.5 mg po daily. |
| M26_Q1 | The features of the American Urologic Association (AUA) symptom score are:  Urinary frequency Urinary hesitancy Weak urinary stream Intermittency Urgency Nocturia Incomplete emptying when urinating Reduced quality of life | The features of the American Urologic Association (AUA) symptom score include:  Urinary frequency Nocturia Incomplete emptying when urinating Urinary hesitancy Weak urinary stream Urinary urgency Urinary intermittence Reduced quality of life | The features of the American Urologic Association (AUA) symptom score include:  Urinary frequency Nocturia Urinary hesitancy Weak urinary stream Urinary urgency Intermittency Incomplete emptying when urinating Reduced quality of life |
| M26_Q2 | Anticholinergics, antihistamines, and opioids can worsen symptoms of benign prostatic hyperplasia. | Anticholinergics, antihistamines, and sympathomimetics can worsen symptoms of benign prostatic hyperplasia. | Anticholinergics, antihistamines, and sympathomimetics can worsen symptoms of benign prostatic hyperplasia. |
| M27_DIFF | The differential diagnosis for this patient's symptoms may include:  Migraine headache with aura Subarachnoid hemorrhage Hypertensive emergency Vertebrobasilar insufficiency Temporal arteritis Non-cardiac chest pain Anxiety disorder Psychogenic nonepileptic seizures. Given the sudden onset and severe nature of the headache, a subarachnoid hemorrhage or a hypertensive emergency should be considered as potential life-threatening causes. The patient's history of palpitations, anxiety, neck swelling, and tachycardia may also suggest a hypertensive emergency. The presence of neck pain, photophobia, and phonophobia may suggest a migraine headache with aura. The transient right-sided arm weakness and altered sensation following the headache may suggest vertebrobasilar insufficiency. The patient's history of temporal pain and neck swelling raises the possibility of temporal arteritis. The patient's nausea and palpitations may suggest non-cardiac chest pain. The patient's anxiety symptoms may indicate an anxiety disorder. The palpitations, neck swelling, anxiety, and fainting spell during a bowel movement may suggest psychogenic nonepileptic seizures. Further evaluation and testing, such as a CT scan and lumbar puncture, may be necessary to determine the underlying cause of the patient's symptoms. | Based on the information provided, the differential diagnosis for the 31-yr-old woman presenting with severe headache and associated symptoms could include:  Migraine headache with aura Subarachnoid hemorrhage Giant cell arteritis Pheochromocytoma Tension headache Cervicogenic headache Hypertensive emergency A thorough evaluation, including a complete history, physical examination, and further tests (such as a non-contrast head CT or lumbar puncture) is necessary to reach a definitive diagnosis and guide appropriate treatment. | Based on the history and physical examination, the following differential diagnoses cannot be excluded in this patient:  Cluster headaches Idiopathic intracranial hypertension Anxiety disorder |
| M27_DIAG | CT scan of the head with IV contrast. | CT scan of the head with IV contrast. | As a language model AI, I don't have the ability to diagnose medical conditions but the following steps are considered essential in emergency medical situations:  Complete blood count (CBC) CT scan of the head with IV contrast |
| M27_DX | Without more information or symptoms, it is not possible to determine the most likely diagnosis. A comprehensive evaluation including a detailed medical history, physical examination, and additional tests may be needed to make an accurate diagnosis. | It is not possible to determine the most likely diagnosis based on the information provided. Further evaluation and diagnostic tests, such as a thorough medical history, neurological examination, and imaging studies, would be necessary to arrive at a definitive diagnosis. | Idiopathic intracranial hypertension |
| M27_MANG | It is not possible to determine the appropriate management without additional information regarding the diagnosis. | Admit the patient to the hospital, MRI of the abdomen and pelvis, ¹²³I-MIBG (iodine-123-labeled metaiodobenzylguanidine) scintigraphy, Ultrasonography of the abdomen, Consult interventional radiology for tumor embolization. | Admit the patient to the hospital Discharge on hydrochlorothiazide 25 mg po once/day plus ramipril 5 mg po once/day Ramipril 5 mg po once/day Propranolol 10 mg po q 6 h Ibuprofen 400 mg tid prn for headache MRI of the abdomen and pelvis Ultrasonography of the abdomen Consult interventional radiology for tumor embolization |
| M27_Q1 | Multiple endocrine neoplasia type 2 (MEN2) and Von Hippel-Lindau (VHL) disease are commonly associated with pheochromocytoma. | Multiple endocrine neoplasia type 2 (MEN2), Von Hippel-Lindau (VHL) disease, Succinate dehydrogenase subunit B-D (SDHB-D) mutations are more commonly associated with pheochromocytoma. | Multiple endocrine neoplasia type 2 (MEN2)  Von Hippel-Lindau (VHL) disease |
| M27_Q2 | NSAIDs, Tricyclic antidepressants, and Monoamine oxidase (MAO) inhibitors are most likely to provoke a hypertensive crisis and therefore are contraindicated in patients with pheochromocytoma. | Monoamine oxidase (MAO) inhibitors and tricyclic antidepressants are most likely to provoke a hypertensive crisis and therefore are contraindicated in patients with pheochromocytoma. | Monoamine oxidase (MAO) inhibitors and Tricyclic antidepressants |
| M27_Q3 | Hypertensive encephalopathy, Left ventricular failure, Pulmonary edema. | Hypertensive encephalopathy, Left ventricular failure, Pulmonary edema. | Hypertensive encephalopathy  Hypoglycemia  Hyponatremia  Left ventricular failure  Pulmonary edema |
| M28_DIFF | At this time, the differential diagnosis should include the following:  Testicular cancer Testicular cyst Spermatocele Testicular torsion Epididymitis/orchitis Hematocele Varicocele | The differential diagnosis for a patient presenting with a hard, nontender mass in the right testis should include:  Testicular cancer Spermatocele Testicular cyst Testicular torsion. | The differential diagnosis for a 28-year-old man with a lump in his right testis should include:  Testicular cancer Testicular cyst Testicular torsion Spermatocele Epididymitis/orchitis |
| M28_DIAG | Scrotal ultrasound and potentially a biopsy of the testis. | The most appropriate next steps in managing this patient would be:  Scrotal ultrasound Consideration of a biopsy of the testis, if necessary based on the ultrasound results. Urinalysis and culture Consideration of a complete blood count (CBC) or sexually transmitted disease (STD) panel, depending on the results of the other tests and the healthcare provider's clinical judgment. | The most appropriate next step in managing this patient would be:  Scrotal ultrasound Biopsy of testis Urinalysis and culture. |
| M28_DX | Testicular cancer is the most likely cause of a solid testicular mass in this patient. | In this patient, the most likely cause of a solid testicular mass is testicular cancer. The presence of a firm, nontender testicular mass that has not gone away after a month is concerning for malignancy, and further diagnostic testing, such as a scrotal ultrasound, and possibly a biopsy, is needed to determine the cause of the mass. However, other causes such as a testicular cyst or a testicular torsion should also be considered and ruled out. | In this patient, the most likely cause of a solid testicular mass is:  Testicular cancer However, a final diagnosis can only be made after a biopsy or other diagnostic tests have been performed. |
| M28_MANG | Urology consultation, CT scan of abdomen and chest, serum tumor markers (alpha-fetoprotein, beta HCG, LDH), and possibly a bone scan. | Urology consultation, CT scan of the abdomen and chest, CBC, serum alfa-fetoprotein, beta HCG, LDH and testosterone levels. | Urology consultation, CT scan of the abdomen and chest, and serum markers (alpha-fetoprotein, beta HCG, LDH) are the most appropriate next steps in managing this patient with a confirmed diagnosis of testicular cancer. |
| M28_Q1 | Testicular cancer is the most common solid cancer in men aged 14-35 years. | Testicular cancer | Testicular cancer |
| M28_Q2 | The most common complaint in patients presenting with a testicular mass is a painless mass on the testis. | Painless mass on testis | Painless mass on testis |
| M28_Q3 | Hydrocele and spermatocele are expected to transilluminate on physical examination. | Hydrocele, spermatocele, and varicocele are expected to transilluminate on physical examination. | Hydrocele and spermatocele are expected to transilluminate on physical examination. |
| M29_DIFF | Based on the history of present illness and physical examination, the differential diagnoses for the 10-year-old boy with constant right-sided scrotal pain should include:  Testicular torsion Orchitis Epididymitis Torsion of the testicular appendage | The differential diagnoses at this time should include:  Testicular torsion Epididymitis Orchitis Torsion of the testicular appendage These conditions are possible causes of scrotal pain in a young male and should be considered based on the patient's presentation. However, a final diagnosis can only be made after further evaluation and imaging studies if necessary. | Epididymitis, Testicular torsion, Testicular fracture, Orchitis. |
| M29_DIAG | Based on the patient's history and physical exam findings, the most appropriate tests to obtain in order to guide diagnosis and management are:  Doppler ultrasonography of the scrotum Urinalysis and culture Additional tests such as a basic metabolic profile (BMP) or complete blood count (CBC) may be considered if there are additional clinical concerns or if the initial test results warrant further investigation. A serum erythrocyte sedimentation rate (ESR) or urethral swab for sexually transmitted disease testing may not be necessary in this case given the patient's history of no recent sexual activity or urethral symptoms. | The most appropriate tests to obtain for this patient based on his history and physical examination are:  Doppler ultrasonography of the scrotum Complete blood count (CBC) Urinalysis and culture Doppler ultrasonography is the most effective imaging modality to confirm or exclude testicular torsion and can be performed quickly and safely in the emergency department. A CBC can help assess for any signs of infection, while a urinalysis and culture can rule out any associated urinary tract infections. Basic metabolic profile and serum erythrocyte sedimentation rate are generally not indicated in this patient's presentation, and a urethral swab for sexually transmitted disease testing is not necessary based on the patient's history of no sexual activity. | Doppler ultrasonography of the scrotum, Urinalysis and culture. |
| M29_DX | Based on the results of the urinalysis and Doppler ultrasonography, the correct diagnosis of this patient's condition is likely:  Testicular cancer (Given the presence of a microcystic lesion adjacent to the right epididymal head and the absence of significant flow within or surrounding the lesion). It is important to note that additional testing and evaluation may be necessary to confirm the diagnosis and rule out other possible causes of scrotal pain. A biopsy of the lesion and further imaging may be required. | Based on the patient's history, physical exam findings, and results of the Doppler ultrasonography of the scrotum, the correct diagnosis at this time is not definitive. The findings of a microcystic lesion on Doppler ultrasonography raise the possibility of a testicular lesion, which could be a testicular cancer or a benign condition such as a testicular cyst, epididymal cyst, or spermatocele. Further evaluation, such as a biopsy or additional imaging studies, may be necessary to confirm or exclude the diagnosis. | Testicular torsion cannot be definitively diagnosed based on the information provided, but the finding of a microcystic lesion on Doppler ultrasonography and the patient's presentation with right-sided scrotal pain suggest it as a possibility. Further evaluation and consultation with a pediatric urologist may be necessary to confirm the diagnosis and guide management. |
| M29_MANG | Admission to the hospital for surgical repair is the most appropriate management of this patient's condition with confirmed torsion of the testicular appendage. The other options are not relevant to the management of torsion of the testicular appendage. | Admission to the hospital for surgical repair and Prescribe oral ibuprofen 300 mg every 6 hours are the most appropriate management options for this patient's condition. | Admission to the hospital for surgical repair. |
| M29_Q1 | The incidence of torsion of an epididymal/testicular appendage is greatest in the prepubertal stage of life. | The incidence of torsion of an epididymal/testicular appendage is greatest in the prepubertal and adolescent stage of life. | The incidence of torsion of an epididymal/testicular appendage is greatest during the prepubertal stage of life. |
| M29_Q2 | The embryologic origin of an epididymal appendage is the mesonephric (wolffian) duct. | The epididymal appendage originates from the mesonephric (wolffian) duct. | The embryologic origin of an epididymal appendage is the mesonephric (wolffian) duct |
| M29_Q3 | The "blue dot sign" is most suggestive of a torsed epididymal/testicular appendage. | The "blue dot sign" is most suggestive of a torsed epididymal/testicular appendage. | The "blue dot sign" is most suggestive of a torsed epididymal/testicular appendage. |
| M30_DIFF | Based on the history and physical examination, the following differential diagnoses cannot be excluded:  Epididymitis or orchitis Testicular appendage torsion Testicular torsion Testicular cancer  It is important to note that the history and physical examination findings suggest that the most likely diagnosis is epididymitis or orchitis, but further testing and imaging is needed to rule out other possibilities such as testicular torsion or cancer. | Based on the given history and physical examination, at this time, the differential diagnoses that cannot be excluded are:  Epididymitis or orchitis Testicular appendage torsion Testicular torsion | Based on the history and physical examination in this patient, the following differential diagnoses cannot be excluded:  Epididymitis or orchitis Testicular appendage torsion Testicular torsion The signs and symptoms of the patient suggest testicular pain and swelling, which could be indicative of various conditions such as epididymitis or orchitis, testicular appendage torsion, or testicular torsion. Further investigation and imaging studies are needed to confirm the diagnosis. |
| M30_DIAG | Given the previous findings, the following are the most appropriate steps to guide diagnosis and treatment:  Urinalysis (UA) and culture Scrotal ultrasonography Sexually transmitted disease (STD) panel  A urinalysis and culture can help to evaluate for any signs of infection in the urinary tract, which can cause epididymitis. Scrotal ultrasonography can help to evaluate the testes and epididymes for any abnormalities such as torsion, infection, or masses. The sexually transmitted disease panel is important because sexually active patients with scrotal pain are at higher risk of acquiring sexually transmitted infections, which can cause epididymitis.  Additional tests such as a complete blood count (CBC) and basic metabolic profile (BMP) may be considered, but are not typically considered first-line tests for evaluating scrotal pain. A biopsy of the testis or an MRI of the pelvis and scrotum may be considered if the initial evaluation is concerning for testicular cancer or other more serious conditions. | Scrotal ultrasonography and possibly a basic metabolic profile (BMP) would be the most appropriate steps to guide diagnosis and treatment in this patient. Urinalysis (UA) and culture may be indicated to rule out related genitourinary issues, and a complete blood count (CBC) may be done to evaluate for any underlying systemic issue contributing to the pain and swelling. Sexually transmitted disease (STD) panel may be done based on the patient's sexual history. Biopsy of the testis is not usually indicated as the first step in the evaluation and management of scrotal pain and swelling, and MRI of pelvis and scrotum is usually not necessary for the initial evaluation of testicular conditions and would be done if other diagnostic modalities failed to provide a clear diagnosis. | Given the previous findings, the following steps would be appropriate to guide diagnosis and treatment:  Urinalysis (UA) and culture Scrotal ultrasonography STD panel (to rule out sexually transmitted infections as a cause) Urinalysis and culture can help evaluate for urinary tract infections, which could cause testicular pain. Scrotal ultrasonography is a non-invasive imaging modality that can help identify the cause of the testicular pain and swelling. An STD panel would be appropriate in a young sexually active male to rule out sexually transmitted infections.  The other tests, such as a CBC and BMP, are not typically used as initial diagnostic tests for testicular pain and swelling and may not provide useful information in this case. A biopsy of the testis would typically only be considered if other diagnostic tests are unable to determine the cause of the symptoms. An MRI of the pelvis and scrotum may also be considered if further evaluation is needed. |
| M30_DX | Based on the results of the testing, the most likely diagnosis in this patient is epididymitis or orchitis. | Based on the results of the testing, the most likely diagnosis in this patient is Epididymitis or Orchitis. The testicular Doppler ultrasonography results show normal echogenicity of the right testicle and epididymitis with increased vascularity compared to the left, suggesting inflammation. The absence of significant findings on urinalysis, urine culture, and the negative gram stain for bacteria support this diagnosis. Testicular torsion, incarcerated inguinal hernia, and testicular appendage torsion can cause similar symptoms, but the ultrasound findings in this case do not support those diagnoses. Testicular cancer is a possible consideration in testicular pain, but the absence of a solid mass and normal echogenicity seen on ultrasound suggest that this is less likely. | Epididymitis or orchitis is the most likely diagnosis in this patient based on the clinical history, physical examination, and test results. The increased vascularity and small, likely reactive right hydrocele on testicular Doppler ultrasonography suggest an acute inflammatory process in the right testicle and epididymis, consistent with epididymitis or orchitis. |
| M30_MANG | Ceftriaxone 250 mg IM and doxycycline 100 mg po twice/day for 10 days Await results of STD panel, then treat accordingly Ibuprofen 400 mg po q 6 h as needed for pain OR Acetaminophen 500 mg po q 6 h as needed for pain Repeat testicular ultrasonography in 1 week Recommend condom usage every time he engages in sexual intercourse | The most appropriate next steps at this time are:  Ceftriaxone 250 mg IM Ibuprofen 400 mg po q 6 h as needed for pain Acetaminophen 500 mg po q 6 h as needed for pain Repeat testicular ultrasonography in 1 week Await results of STD panel, then treat accordingly Recommend condom usage every time he engages in sexual intercourse | Ibuprofen 400 mg po q 6 h as needed for pain  Await results of STD panel, then treat accordingly  Repeat testicular ultrasonography in 1 week  Recommend condom usage every time he engages in sexual intercourse |
| M30_Q1 | Chlamydia trachomatis, Neisseria gonorrhoeae. | Chlamydia trachomatis and Neisseria gonorrhoeae are common organisms associated with epididymitis in men under 35 years old. | Chlamydia trachomatis and Neisseria gonorrhoeae are common organisms associated with epididymitis in men <35 years old. |
| M30_Q2 | Cytomegalovirus and tuberculosis are more common causes of epididymitis or orchitis in immunocompromised patients. | Cytomegalovirus and tuberculosis are more common causes of epididymitis or orchitis among immunocompromised patients. | Cytomegalovirus, tuberculosis and blastomycosis are more common causes of epididymitis or orchitis among immunocompromised patients. |
| M30_Q3 | Infertility and testicular atrophy can develop after mumps orchitis. | Infertility and testicular atrophy commonly develop after mumps orchitis. | Infertility and testicular atrophy are the conditions that commonly develop after mumps orchitis. |
| M31_DIFF | Based on the given history and physical examination, the following causes of the patient's exertional dyspnea CANNOT be excluded:  Acute decompensated heart failure (HF) COPD exacerbation Pneumonia Pneumothorax Note: Further diagnostic tests, such as a chest x-ray, electrocardiogram (ECG), and laboratory tests, are necessary to establish a definite diagnosis. | Based on the history and physical examination, the following causes of the patient's exertional dyspnea cannot be excluded:  Acute decompensated heart failure (HF) COPD exacerbation Pneumonia Pulmonary embolism (PE) | Based on the history and physical examination, none of the following can be excluded as causes of the patient's exertional dyspnea:  Acute decompensated heart failure (HF) COPD exacerbation Pneumonia Pulmonary embolism |
| M31_DIAG1 | Electrocardiogram (ECG), Serum troponin level, Chest x-ray (CXR), and Transthoracic echocardiography with Doppler ultrasonography (TTE) are the most appropriate steps to help establish a diagnosis. | It depends on the specific case and the suspected cause(s) of exertional dyspnea. However, some commonly used diagnostic tools for this condition include:  Electrocardiogram (ECG) Chest x-ray (CXR) Arterial blood gas Transthoracic echocardiography with Doppler ultrasonography (TTE) Serum troponin level Serum brain natriuretic peptide (BNP) level Additional testing such as CT scan of the chest, pulmonary function testing, cardiac MRI, and others may be done based on the results of the initial tests and the clinician's suspicion for the underlying cause. | It depends on the individual's symptoms and suspected medical condition. However, some of the most commonly used diagnostic tests to help establish a diagnosis include:  Electrocardiogram (ECG) Chest x-ray (CXR) Serum troponin level Transthoracic echocardiography with Doppler ultrasonography (TTE) CT scan of the chest Cardiac MRI Serum brain natriuretic peptide (BNP) level Arterial blood gas Pulmonary function testing. |
| M31_DX | The most likely diagnosis based on the results provided is Acute decompensated heart failure. The elevated BNP level, presence of perihilar opacities and small bilateral pleural effusions on chest x-ray, and restrictive LV diastolic pattern with biatrial enlargement on transthoracic echocardiography are all consistent with this diagnosis. The normal serum troponin level rules out Acute Myocardial Infarction and the absence of pericardial thickening and pericardial calcifications on chest x-ray and transthoracic echocardiography suggests against Cardiac tamponade or Constrictive pericarditis. | Constrictive pericarditis is the most likely diagnosis based on the test results. | The most likely diagnosis based on the results provided is Acute decompensated heart failure. The high BNP level, biatrial enlargement, restrictive LV diastolic pattern, inspiratory decrease of 5% in mitral diastolic flow velocity and perihilar opacities on the chest x-ray all suggest signs of heart failure. Additionally, the normal left ventricular size and function with severe concentric left ventricular hypertrophy is consistent with heart failure. The normal troponin level rules out acute myocardial infarction (MI) and the normal pericardial appearance on TTE rules out cardiac tamponade and constrictive pericarditis. |
| M31_DIAG2 | Cardiac catheterization and Transesophageal echocardiogram (TEE) are the most appropriate steps for guiding diagnosis and treatment of ADHF, as they can provide further information about the underlying cardiac conditions, such as the degree of stenosis or occlusion in the coronary arteries and the presence of valvular disease. Dobutamine stress echocardiography and exercise stress test may be useful to assess the functional status of the heart, but are not considered primary diagnostic tools for ADHF. | At this time, the most appropriate steps for guiding diagnosis and treatment of the confirmed ADHF would be:  Cardiac catheterization Transesophageal echocardiogram (TEE) | Cardiac catheterization and Transesophageal echocardiogram (TEE) are the most appropriate steps for guiding diagnosis and treatment in this case of confirmed acute decompensated heart failure (ADHF). |
| M31_Q1 | Based on the results of the cardiac catheterization, the most likely etiology of the patient's acute decompensated heart failure (ADHF) is restrictive cardiomyopathy, as indicated by the prominence of the y descent on the RA waveform and the concordance in LV and RV pressure tracings with respiration. The LVEDP of 18 mm Hg and the PCWP of 17 mm Hg support this diagnosis. | Based on the results of the cardiac catheterization, the most likely etiology of the patient's ADHF is restrictive cardiomyopathy. The findings of a restrictive LV diastolic pattern, prominent y descent on the RA waveform, and concordance in LV and RV pressure tracings with respiration suggest restrictive cardiomyopathy, which is characterized by limited filling of the ventricles during diastole. | Based on the results of the cardiac catheterization, the most likely etiology of the patient's ADHF is restrictive cardiomyopathy. The presence of a prominent y descent in the right atrial waveform, concordance between LV and RV pressure tracings with respiration, and the findings of a restrictive LV diastolic pattern with elevated LVEDP (left ventricular end-diastolic pressure) and PCWP (pulmonary capillary wedge pressure) on TTE (transthoracic echocardiogram) are consistent with restrictive cardiomyopathy. |
| M31_DIAG3 | Endomyocardial biopsy and Serum ferritin level are the most appropriate steps to determine the etiology of the patient's restrictive cardiomyopathy. Endomyocardial biopsy can provide tissue samples for histologic and immunologic examination to identify specific causes of restrictive cardiomyopathy, such as infiltrative diseases or storage disorders. Serum ferritin level can help determine if iron overload, a cause of restrictive cardiomyopathy, is present. A complete blood count (CBC) with differential and fat pad biopsy are not typically indicated for determining the etiology of restrictive cardiomyopathy. | A diagnosis of restrictive cardiomyopathy has been established based on the test results. To determine the etiology of this patient's restrictive cardiomyopathy, the following steps would be appropriate:  Endomyocardial biopsy Fat pad biopsy Serum ferritin level Complete blood count (CBC) with differential | The most appropriate steps to determine the etiology of this patient's restrictive cardiomyopathy are:  Endomyocardial biopsy Fat pad biopsy Endomyocardial biopsy is considered the gold standard for diagnosing many causes of restrictive cardiomyopathy, including infiltrative diseases, such as amyloidosis. Fat pad biopsy can be used to diagnose amyloidosis.  A complete blood count (CBC) with differential and serum ferritin level are not specific tests for restrictive cardiomyopathy and may not provide any useful information in this context. |
| M31_Q2 | The most likely etiology of the patient's restrictive cardiomyopathy is Restrictive cardiomyopathy due to cardiac amyloidosis based on the fat pad biopsy results which showed amorphous extracellular pink deposits with hematoxylin and eosin stain and apple-green birefringence under polarized microscopy on Congo red staining. | Based on the result of the fat pad biopsy, the most likely etiology of the patient's restrictive cardiomyopathy is Restrictive cardiomyopathy due to cardiac amyloidosis. The presence of amorphous extracellular pink deposits with apple-green birefringence under polarized microscopy on Congo red staining supports the diagnosis of cardiac amyloidosis. | Restrictive cardiomyopathy due to cardiac amyloidosis |
| M31_MANG | Based on the current information, the most appropriate management options for this patient are:  Admit to a monitored telemetry inpatient unit for further treatment Antiplasma cell therapy Sodium restriction Fluid restriction Amyloid typing | Admit to a monitored telemetry inpatient unit for further treatment Furosemide 40 mg IV bolus followed by 10 mg/hour furosemide infusion 2-g sodium restriction 1-L fluid restriction Amyloid typing | Admit to a monitored telemetry inpatient unit for further treatment, Furosemide 40 mg IV bolus followed by 10 mg/hour furosemide infusion, 2-g sodium restriction, 1-L fluid restriction, Lisinopril 20 mg orally daily, Metoprolol 50 mg orally daily, and Amyloid typing. |
| M31_Q3 | ACE inhibitors, Aldosterone antagonists, Angiotensin II receptor blockers, Beta blockers, and some studies suggest that Digoxin may decrease mortality in patients with heart failure with reduced ejection fraction. | ACE inhibitors, Aldosterone antagonists, Angiotensin II receptor blockers, and Beta blockers have been shown to decrease mortality in patients with heart failure with reduced ejection fraction. | Angiotensin-converting enzyme (ACE) inhibitors, Aldosterone antagonists, Angiotensin receptor blockers (ARBs), Beta blockers, and Ivabradine (a selective sinus node inhibitor) have been shown to decrease mortality in patients with heart failure with reduced ejection fraction. |
| M31_Q4 | The following factors suggest candidacy for cardiac resynchronization therapy in heart failure:  Left ventricular ejection fraction <35% Nonspecific intraventricular conduction delay with mildly prolonged QRS (QRS >130 msec) New York Heart Association (NYHA) class III or IV symptoms | The following factors suggest candidacy for cardiac resynchronization therapy in heart failure:  Left ventricular ejection fraction <35% Nonspecific intraventricular conduction delay with mildly prolonged QRS (QRS >130 msec) NYHA class III or IV symptoms Sinus rhythm. | The following factors suggest candidacy for cardiac resynchronization therapy in heart failure:  Left ventricular ejection fraction <35% Nonspecific intraventricular conduction delay with mildly prolonged QRS (QRS >130 msec) New York Heart Association (NYHA) class III or IV symptoms |
| M31_Q5 | Common findings in patients with amyloid cardiomyopathy include:  Dilated cardiomyopathy Diastolic dysfunction Heart failure Arrhythmias Heart block. | Heart failure, diastolic dysfunction, and arrhythmias are common findings in patients with amyloid cardiomyopathy. | Common findings in patients with amyloid cardiomyopathy include:  Diastolic dysfunction Heart failure Arrhythmias Heart block |
| M31_Q6 | The following may be effective in the treatment of light-chain (AL) cardiac amyloidosis:  Antiplasma cell therapy Autologous stem cell transplantation Heart transplantation | Light-chain (AL) cardiac amyloidosis is a life-threatening disease, and there are several treatment options that can be considered, although no cure is currently available. Some of the effective treatment options for AL cardiac amyloidosis include:  Antiplasma cell therapy: Chemotherapy and stem cell transplantation have shown some promise in treating AL amyloidosis.  Autologous stem cell transplantation: This procedure involves removing stem cells from the patient's own blood, treating the cells to eliminate the plasma cells producing the abnormal protein, and then reinfusing the cells back into the patient's body.  Heart transplantation: Heart transplantation may be considered as a treatment option for patients with end-stage heart failure due to AL amyloidosis, but it is a complex procedure with a high risk of morbidity and mortality.  It's important to note that treatment decisions should be made in collaboration with a multidisciplinary team of healthcare professionals, considering the individual patient's overall health status, comorbidities, and goals of care. | Antiplasma cell therapy, autologous stem cell transplantation, and heart transplantation may be effective in the treatment of light-chain (AL) cardiac amyloidosis. |
| M32_DIFF | At this time, the differential diagnoses that cannot be excluded based on history and physical examination in this patient are:  Acute coronary syndrome/ST segment elevation myocardial infarction (STEMI) Aortic dissection Dehydration Dysrhythmia Neurogenic shock Pericardial effusion with cardiac tamponade Pulmonary embolism (PE) Septic shock Stroke/transient ischemic attack (TIA) Vasovagal syncope. | Based on the history and physical examination, the following differential diagnoses cannot be excluded:  Acute coronary syndrome/STEMI Aortic dissection Pulmonary embolism (PE) Pericardial effusion with cardiac tamponade Septic shock Stroke/transient ischemic attack (TIA) Vasovagal syncope | Acute coronary syndrome/ST segment elevation myocardial infarction (STEMI), Aortic dissection, Dehydration, Dysrhythmia, Pericardial effusion with cardiac tamponade, Pulmonary embolism (PE), Septic shock, Stroke/transient ischemic attack (TIA), and Vasovagal syncope cannot be excluded based on history and physical examination in this patient. |
| M32_MANG1 | Bedside glucose measurement, BP measurement every 5 min, pulse oximetry, cardiac monitoring, oxygen 15 L by non-rebreather mask, and 0.9% saline solution, 1 L IV bolus. | BP measurement every 5 min, Cardiac monitoring, Oxygen 15 L by nonrebreather mask, 0.9% saline solution, 1 L IV bolus, and Dextrose 50 g IV. | Bedside glucose measurement, BP measurement every 5 min, pulse oximetry, cardiac monitoring, and oxygen 15 L by nonrebreather mask, 0.9% saline solution, 1 L IV bolus are essential to do immediately in this patient. |
| M32_DIAG | ECG and Chest x-ray at bedside. | ECG and Chest x-ray at bedside | ECG and Chest x-ray at bedside. |
| M32_DX | The most likely diagnosis based on the given information is cardiac tamponade. | Based on the recent results, it is difficult to definitively determine the most likely diagnosis. Sinus tachycardia and a slightly enlarged cardiac silhouette on chest x-ray suggest possible cardiac involvement, but the lack of ST-T wave abnormalities and electrical alternans on the ECG make acute coronary syndrome less likely. Normal voltage on the ECG makes cardiac tamponade less likely. The normal ECG findings also make dysrhythmia less likely. The normal chest x-ray findings do not support pulmonary embolism. Further testing and evaluation may be needed to reach a more definitive diagnosis. | Based on the information provided, it is most likely that the patient is suffering from Sinus Tachycardia. Sinus Tachycardia is a condition where the heart beats faster than normal due to an underlying cause such as anxiety, fever, dehydration, and more. The ECG results show sinus tachycardia, normal voltage, and no ST-T wave abnormalities or electrical alternans, which supports this diagnosis. However, further testing may be necessary to rule out other possible conditions and arrive at a definitive diagnosis. |
| M32_MANG2 | Consult cardiothoracic surgeon and Pericardiocentesis, done in an operating room. | Consult cardiothoracic surgeon, Pericardiocentesis, done in an operating room. | Consult cardiothoracic surgeon and pericardiocentesis, done in an operating room. |
| M32_Q1 | Pulsus paradoxus is often associated with Cardiac tamponade. | Pulsus paradoxus is often associated with the following conditions:  Cardiac tamponade Asthma exacerbation Pulsus paradoxus refers to a significant drop in systolic blood pressure during inspiration that occurs in certain medical conditions, including cardiac tamponade and severe asthma. However, it is important to note that the presence of pulsus paradoxus does not confirm the diagnosis, but it can be supportive evidence in the context of other findings. | Pulsus paradoxus is often associated with Cardiac tamponade. |
| M32_Q2 | Hypotension, Jugular venous distension, Muffled heart sounds, and Tachycardia are commonly associated with cardiac tamponade. | Hypotension, jugular venous distention, muffled heart sounds, and tachycardia are commonly associated with cardiac tamponade. | Hypotension, jugular venous distension, muffled heart sounds, and tachycardia are commonly associated with cardiac tamponade. |
| M32_Q3 | Electrical alternans (beat-to-beat QRS amplitude variability), S₁, Q₃, and T₃ pattern. | Electrical alternans (beat-to-beat QRS amplitude variability) and Tachycardia are classically associated with pericardial effusion and cardiac tamponade. | The ECG findings classically associated with pericardial effusion and cardiac tamponade are:  Electrical alternans (beat-to-beat QRS amplitude variability) S₁, Q₃, and T₃ Tachycardia |
| M33_DIFF | Onset of labor, Placenta previa, and Placental abruption should be included in the diagnoses. | Onset of labor, Placental abruption, Miscarriage, Placenta previa, Vasa previa | Based on the information provided, the possible diagnoses at this time include:  Miscarriage Onset of labor Placenta previa Placental abruption Vasa previa It is important to keep in mind that the information provided is limited and additional diagnostic tests may be necessary to confirm a diagnosis. |
| M33_DIAG1 | Complete blood count (CBC), Fetal ultrasonography, External fetal monitoring with tocometry, and Crossmatch 2 units of packed red blood cells are the most appropriate initial diagnostic studies. | The most appropriate initial diagnostic studies for this patient would be:  Complete blood count (CBC) Blood type and screen External fetal monitoring with tocometry Fetal ultrasonography Digital vaginal exam Kleihauer-Betke test These studies would help assess the patient's bleeding status and rule out any potential life-threatening conditions, such as a placental abruption or vasa previa, as well as evaluate the fetus's well-being and rule out any other possible causes of vaginal bleeding, such as cervical or endometrial abnormalities. | The most appropriate initial diagnostic studies at this time would include:  Complete blood count (CBC) Fetal ultrasonography External fetal monitoring with tocometry Fetal fibronectin These tests can help determine the cause of the vaginal bleeding, the well-being of the fetus, and the potential for preterm labor. Additional diagnostic studies may be necessary based on the results of these tests. |
| M33_DX | Abnormal fetal status Placental abruption Preterm labor | Abnormal fetal status, Placental abruption, Preterm labor | Abnormal fetal status, Placental abruption, Preterm labor |
| M33_MANG | Immediate cesarean section Transfuse 2 units packed red blood cells (PRBCs) | Admit for fetal monitoring and delivery at 37 weeks Immediate cesarean section Transfuse 2 units packed red blood cells (PRBCs) | Immediate cesarean section, Transfuse 2 units packed red blood cells (PRBCs) |
| M33_Q1 | Chronic hypertension, history of placental abruption, tobacco use, and older maternal age are risk factors for placental abruption. | Chronic hypertension, Cocaine use, History of placental abruption, Nulliparity, Tobacco use, Younger maternal age. | Chronic hypertension, History of placental abruption, Cocaine use, Tobacco use, and Abdominal trauma. |
| M33_Q2 | A digital vaginal exam is contraindicated in the following causes of vaginal bleeding in late pregnancy:  Placenta previa  Abruptio placentae | Digital vaginal exam is contraindicated in Abruptio placentae and Placenta previa. | A digital vaginal exam is contraindicated in the following causes of vaginal bleeding in late pregnancy:  Abruptio placentae Placenta previa Uterine rupture |
| M34_Q1 | Cranial nerve III, IV and VI. | Cranial nerve III, IV, and VI | Cranial nerve III, IV and VI |
| M34_DIFF | At this time, based on the limited information provided, none of the differential diagnoses can be excluded. Further testing and evaluation would be necessary to determine the correct diagnosis. | Without additional information, it is impossible to determine which differential diagnoses cannot be excluded based on history and physical examination in this patient. Further testing, such as imaging studies and laboratory tests, would be necessary to arrive at a definitive diagnosis. | Without any additional information about the patient, it is not possible to exclude any of the differential diagnoses listed. |
| M34_DIAG | Midnight serum cortisol, serum acetylcholine receptor antibody levels, serum TSH, and MRI of the cranium. | As a language model, I can't provide a definitive answer without further context. The most appropriate next steps would depend on the specific symptoms and signs present in the patient. A healthcare professional would need to evaluate the patient's complete medical history and physical examination to determine which tests are necessary. However, some tests that are commonly used to evaluate patients with a wide range of symptoms and conditions include: CBC, electrolyte concentrations, liver function studies (LFTs), blood glucose, TSH, FT4, and MRI of the cranium. | It depends on the specific symptoms and history of the patient. In general, the following tests might be considered:  Blood glucose: to check for uncontrolled diabetes Midnight serum cortisol: to check for Cushing syndrome Serum free T4 (FT4), serum TSH: to check for hypothyroidism Serum acetylcholine receptor antibody levels: to check for myasthenia gravis Serum electrolyte concentrations: to check for electrolyte imbalances Serum lactate dehydrogenase (LDH), serum liver function studies (LFT), complete blood count (CBC): to assess for muscle inflammation or damage MRI of the cranium: to evaluate for a brain tumor Note: These recommendations are not exhaustive and the most appropriate next steps would depend on the individual case and the clinical judgment of the physician |
| M34_DX1 | Based on the results provided, the single diagnosis that best accounts for the patient's findings is Hypothyroidism. The patient has a low FT4 level and a high TSH level, which are consistent with hypothyroidism. | Hypothyroidism | Based on the results, the single diagnosis that best accounts for the patient's findings is Diabetes, uncontrolled. The high glucose level of 17.0 mmol/L (306 mg/dL) and negative capillary blood ketones suggest uncontrolled diabetes, while the normal LFT, TSH, and FT4 levels do not support hypothyroidism, Cushing syndrome, or hypokalemia as the primary diagnosis. |
| M34_MANG1 | Chest x-ray, Inferior petrosal sinus sampling for serum ACTH and cortisol levels, Serum ACTH level, Serum corticotropin-releasing hormone (CRH) test | The most appropriate next steps for diagnosis and further evaluation of Cushing syndrome based on the results of the tests are:  Inferior petrosal sinus sampling for serum ACTH and cortisol levels Oral glucose tolerance test Chest x-ray. | Chest x-ray, Inferior petrosal sinus sampling for serum ACTH and cortisol levels, Oral glucose tolerance test, Serum ACTH level, Serum corticotropin-releasing hormone (CRH) test, Serum LH level, Serum prolactin level, Serum testosterone level. |
| M34_DX2 | Based on the laboratory results, the most likely cause of Cushing syndrome in this patient is Ectopic ACTH secretion. | Based on the results, the most likely cause of Cushing syndrome in this patient is Ectopic ACTH secretion. | The most likely cause of Cushing syndrome in this patient is Ectopic ACTH secretion, based on the elevated levels of ACTH and cortisol in the blood and an unremarkable chest x-ray. |
| M34_MANG2 | The appropriate management steps for this patient with pituitary ACTH-dependent Cushing syndrome are:  Transsphenoidal adenomectomy Pegvisomant 40 mg once a day Watch and wait, with regular monitoring of serum cortisol levels and repeat MRI of the pituitary yearly | The most appropriate management steps for this patient with confirmed pituitary ACTH-dependent Cushing syndrome are:  Transsphenoidal adenomectomy Pegvisomant 40 mg once a day Watch and wait, with regular monitoring of serum cortisol levels and repeat MRI of the pituitary yearly | The most appropriate management steps for pituitary ACTH-dependent Cushing syndrome include:  Transsphenoidal adenomectomy Pegvisomant 40 mg once a day Watch and wait, with regular monitoring of serum cortisol levels and repeat MRI of the pituitary yearly |
| M34_Q3 | III, IV, or/and VI cranial nerve palsy Bitemporal hemianopia due to an optic chiasm compression Pituitary tumor apoplexy Deep vein thrombosis | The complications of Cushing disease are:  Bitemporal hemianopia due to an optic chiasm compression Pituitary tumor apoplexy III, IV, or/and VI cranial nerve palsy | III, IV, or/and VI cranial nerve palsy  Bitemporal hemianopia due to an optic chiasm compression  Pituitary tumor apoplexy  Deep vein thrombosis |
| M34_Q4 | Breast cancer  Small-cell lung carcinoma  Neuroendocrine tumor | Breast cancer  Endometrial carcinoma  Neuroendocrine tumor  Small-cell lung carcinoma | Breast cancer, small-cell lung carcinoma, neuroendocrine tumor, and medullary thyroid carcinoma can be possible causes of an ectopic ACTH-dependent Cushing syndrome. |
| M34_Q5 | At least two 24-hour collections of urinary free cortisol  Overnight dexamethasone suppression test  Midnight sleeping serum or salivary cortisol | Midnight sleeping serum or salivary cortisol  Overnight dexamethasone suppression test | The first-line (screening) tests for confirmation of endogenous hypercortisolism in patients with suspected Cushing syndrome are:  24-hour urinary free cortisol collection Overnight dexamethasone suppression test Midnight serum or salivary cortisol test |
| M34_MANG3 | Bilateral adrenalectomy  Transsphenoidal pituitary adenomectomy  Pegvisomant  Ketoconazole or metyrapone  Pituitary radiotherapy | Bilateral adrenalectomy Transsphenoidal pituitary adenomectomy Pegvisomont Ketoconazole or Metyrapone Pituitary radiotherapy | Bilateral adrenalectomy Metyrapone or ketoconazole Transsphenoidal pituitary adenomectomy Pegvisomant Pituitary radiotherapy |
| M35_DIFF | Based on the history and physical examination, the following diagnoses cannot be excluded:  Adrenal insufficiency Anemia Cancer Gastroenteritis GI bleeding Hypothyroidism Liver failure Renal failure Sepsis. | Adrenal insufficiency, Anemia, Cancer, Gastroenteritis, GI bleeding, Heart failure, Hyperthyroidism, Hypothyroidism, Liver failure, Renal failure, Sepsis | Based on the history and physical examination, the following diagnoses cannot be excluded:  Adrenal insufficiency Anemia Cancer GI bleeding Heart failure Hyperthyroidism Hypothyroidism Liver failure Renal failure Sepsis |
| M35_DIAG | The appropriate steps for guiding diagnosis and treatment depend on the individual patient's symptoms and medical history. However, some commonly used tests include:  Complete blood count (CBC) Serum electrolyte concentrations, creatinine, and urea Serum liver function studies (LFTs) and amylase Serum calcium Serum troponin Chest x-ray CT scan of the abdomen Echocardiography Bedside blood glucose (in specific cases where the patient presents symptoms of hypoglycemia) It's important to note that these tests may not be appropriate in every case, and a healthcare provider should evaluate each patient on a case-by-case basis to determine the best course of action. | Serum electrolyte concentrations, creatinine, and urea Complete blood count (CBC) Chest x-ray Serum liver function studies (LFTs) Venous blood gas for pH, bicarbonates, and lactate CT scan of the abdomen Serum troponin Serum calcium Echocardiography Urine dipstick | It is not possible to determine the most appropriate steps for guiding diagnosis and treatment without more information about the patient's symptoms and medical history. The appropriate tests will depend on the individual case and the healthcare provider's clinical judgement. However, some common tests that may be useful in various scenarios include a CBC, serum electrolyte concentrations, creatinine and urea, serum TSH, serum free T4 (FT4), serum liver function studies (LFTs) and amylase, serum calcium, and a urine dipstick. |
| M35_DX1 | Based on the results, it is most likely a new presentation of type 1 diabetes. | Based on the results, the most likely diagnosis is cortisol insufficiency complicated with an acute kidney injury. | Hyperthyroidism is the most likely diagnosis. |
| M35_MANG1 | Serum ACTH level Overnight dexamethasone suppression test Short Synacthen test CT/MRI of the adrenal glands MRI of the pituitary gland | Serum ACTH level, Short Synacthen test, CT/MRI of the adrenal glands, and MRI of the pituitary gland. | Serum ACTH level, Overnight dexamethasone suppression test, CT/MRI of the adrenal glands, MRI of the pituitary gland, and Screening for tuberculosis are the most appropriate next steps to evaluate and manage the patient's cortisol deficiency. |
| M35_DX2 | The most likely cause of this patient's adrenal insufficiency is bilateral subacute adrenal hemorrhage, as reported in the CT scan of the adrenal glands. | The most likely cause of this patient's adrenal insufficiency is bilateral adrenal hemorrhage following major surgery, as indicated by the CT scan result of bilateral subacute adrenal hemorrhage. | Bilateral adrenal hemorrhage following major surgery. |
| M35_MANG2 | Hydrocortisone 50 mg IV every 6 hours 0.9% saline IV 250 mL/hr Oral hydrocortisone (10 mg on waking up, 5 mg at midday and 5 mg around 5 PM) orally Fludrocortisone 50 mcg once a day orally | The appropriate management steps for this patient with a diagnosis of Addisonian crisis due to bilateral adrenal hemorrhage are:  Hydrocortisone 50 mg IV every 6 hours 0.9% saline IV 250 mL/hr Fludrocortisone 50 mcg once a day orally Referral to an endocrine surgeon for consideration of treatment options, including surgical management Regular monitoring of serum cortisol levels and repeat MRI of the adrenal glands in 6 months. | The appropriate management steps for this patient include:  Hydrocortisone 50 mg IV every 6 hours Fludrocortisone 50 mcg once a day orally 0.9% saline IV 250 mL/hr Regular monitoring of serum cortisol levels and repeat MRI of the adrenal glands in 6 months Transition to oral hydrocortisone (10 mg on waking up, 5 mg at midday and 5 mg around 5 PM) when the patient is stable |
| M35_Q1 | The causes of primary adrenal insufficiency include:  Autoimmune adrenalitis Adrenal tuberculosis Congenital adrenal hyperplasia Bilateral adrenalectomy Bilateral adrenal hemorrhage Amyloidosis or hemochromatosis Adrenoleucodystrophy Carney complex Bilateral metastases to the adrenal glands Histoplasmosis Sarcoidosis | The causes of primary adrenal insufficiency include:  Autoimmune adrenalitis Adrenal tuberculosis Congenital adrenal hyperplasia Adrenoleucodystrophy Amyloidosis or hemochromatosis Bilateral adrenal hemorrhage Bilateral metastases to the adrenal glands Carney complex Histoplasmosis Pendred syndrome Sarcoidosis | Adrenal tuberculosis, autoimmune adrenalitis, bilateral adrenalectomy, bilateral adrenal hemorrhage, bilateral metastases to the adrenal glands, congenital adrenal hyperplasia, histoplasmosis, and sarcoidosis are causes of primary adrenal insufficiency. |
| M35_Q2 | Hyponatremia, hyperkalemia, and metabolic acidosis are biochemical abnormalities of adrenal deficiency. | Hyponatremia Hypoglycemia Hyperkalemia Metabolic acidosis | Hyponatremia, Hyperkalemia, and metabolic acidosis are biochemical abnormalities of adrenal deficiency. |
| M35_Q3 | An Addisonian crisis can be provoked by:  Rapid withdrawal of long-term prednisolone Strenuous exercise; eg, running a marathon Surgery with a general anesthetic Meningococcal septicemia Pneumonia Gastroenteritis Extensive burn injury CT scan with contrast Significant weight loss Uncomplicated tooth extraction | An Addisonian crisis can be provoked by:  Rapid withdrawal of long-term prednisolone Strenuous exercise; eg, running a marathon Surgery with a general anesthetic Gastroenteritis Pneumonia Meningococcal septicemia Significant weight loss Extensive burn injury CT scan with contrast | An Addisonian crisis can be provoked by:  Rapid withdrawal of long-term prednisolone Significant weight loss Strenuous exercise; eg, running a marathon Surgery with a general anesthetic Gastroenteritis Pneumonia Meningococcal septicemia Extensive burn injury CT scan with contrast |
| M35_Q4 | Hyperpigmentation, postural dizziness, salt craving, syncope/cardiovascular collapse, tiredness, weight loss are symptoms and signs of primary adrenal insufficiency. | Hyperpigmentation, postural dizziness, salt craving, tiredness, weight loss. | Hyperpigmentation, Salt craving, Tiredness, Weight loss, Nausea and vomiting, Postural dizziness, Syncope/cardiovascular collapse, Abdominal pain, Bone fractures. |
| M36_DIFF | The differential diagnoses at this time may include:  Hyperthyroidism Malabsorption Occult malignancy Tuberculosis | Based on the patient's symptoms, the differential diagnoses that should be considered at this time include:  Hyperthyroidism (presenting with symptoms of weight loss, palpitations, sweating, irritability, and tremors) Malabsorption (presenting with weight loss, loose stools, and possibly fatigue) Occult malignancy (presenting with weight loss, fatigue, and other non-specific symptoms) Tuberculosis (presenting with weight loss, fatigue, and other non-specific symptoms) It is important to note that further evaluation and laboratory tests would be necessary to determine the exact diagnosis. | Hyperthyroidism, Malabsorption, Occult malignancy, Tuberculosis. |
| M36_DIAG | The most appropriate studies to obtain to guide diagnosis are:  Complete blood count (CBC) Serum glucose level Chest x-ray ECG Serum TSH, free T4, and free T3 levels Serum electrolytes, BUN, and creatinine levels | Serum TSH, free T4, and free T3 levels, Complete blood count (CBC), Serum electrolytes, BUN, and creatinine levels, Chest x-ray, ECG. | Serum TSH, free T4, and free T3 levels, Chest x-ray, ECG, Complete blood count (CBC), Serum electrolytes, BUN, and creatinine levels. |
| M36_DX | The most likely diagnosis for this patient is Primary hyperthyroidism based on the results of the serum TSH, free T4, and free T3 levels. The low TSH and high FT4 and FT3 levels suggest overproduction of thyroid hormones by the thyroid gland, which is a characteristic of hyperthyroidism. The sinus tachycardia seen on the ECG is also consistent with hyperthyroidism. The other test results, including the normal chest x-ray and normal electrolyte levels, do not suggest any other specific diagnosis. | The most likely diagnosis for this patient is primary hyperthyroidism. The test results show low TSH levels (<0.01 mU/L) and elevated FT4 (4.2 ng/dL) and FT3 (15.5 pg/mL) levels, which are consistent with hyperthyroidism. Sinus tachycardia (110 bpm) seen on ECG is also a common finding in hyperthyroidism. The chest x-ray and CBC results are normal, which does not support other diagnoses like malabsorption or endocarditis. The patient's random glucose level (126 mg/dL) does not meet the diagnostic criteria for diabetes. The absence of significant findings on history and physical examination suggests that drug effects are unlikely. | Primary hyperthyroidism |
| M36_MANG1 | The most appropriate next steps to further evaluate the patient's hyperthyroidism are:  Serum TSH receptor antibodies Technetium-99m thyroid uptake scan | The most appropriate next steps to further evaluate the patient's hyperthyroidism would be a technetium-99m thyroid uptake scan and serum TSH receptor antibodies. | Serum TSH receptor antibodies and Technetium-99m thyroid uptake scan are the most appropriate next steps to further evaluate the patient's hyperthyroidism. |
| M36_DX2 | Based on the positive result of serum TSH receptor antibodies, the most likely cause of hyperthyroidism in this patient is Graves disease. | Based on the positive result for serum TSH receptor antibodies, the most likely cause of hyperthyroidism in this patient is Graves disease. | Given the presence of positive TSH receptor antibodies, the most likely cause of hyperthyroidism in this patient is Graves disease. |
| M36_MANG2 | Based on the results and the patient's diagnosis of Graves' disease, the most appropriate management steps for this patient would be:  Methimazole 20 mg orally twice a day Propylthiouracil 150 mg orally 3 times a day Levothyroxine 100 mcg orally once a day Propranolol 40 mg orally 3 times a day Hypromellose eye drops 4 times a day and Lacrilube eye ointment at night Smoking cessation program Note: The other options listed are not appropriate for this patient's diagnosis and management. | The most appropriate management steps for this patient with hyperthyroidism include:  Propylthiouracil 150 mg orally 3 times a day Methimazole 20 mg orally twice a day Levothyroxine 100 mcg orally once a day Propranolol 40 mg orally 3 times a day Hypromellose eye drops 4 times a day and Lacrilube eye ointment at night Smoking cessation program | The most appropriate management steps for this patient are:  Methimazole 20 mg orally twice a day Propylthiouracil 150 mg orally 3 times a day Levothyroxine 100 mcg orally once a day Hypromellose eye drops 4 times a day and Lacrilube eye ointment at night Smoking cessation program Radioactive iodine-131 400 MBq (as a possible long-term management option) |
| M36_Q1 | Graves disease, Amiodarone-induced thyrotoxicosis, Iodine-induced thyrotoxicosis, Subacute thyroiditis, Toxic multinodular goiter, and TSH-producing pituitary adenoma are all causes of hyperthyroidism. | The causes of hyperthyroidism are:  Graves disease Toxic multinodular goiter Subacute thyroiditis Iodine-induced thyrotoxicosis Amiodarone-induced thyrotoxicosis Factitious toxicosis Postpartum thyroiditis Struma ovari Thyroid cyst. | The following are causes of hyperthyroidism:  Graves disease Toxic multinodular goiter Iodine-induced thyrotoxicosis Subacute thyroiditis Amiodarone-induced thyrotoxicosis Factitious toxicosis Postpartum thyroiditis Thyroid cyst TSH-producing pituitary adenoma |
| M36_Q2 | The potential complications of Graves disease include:  Thyroid eye disease Atrial fibrillation Thyroid storm Pretibial myxedema | Graves disease can result in the following potential complications:  Thyroid eye disease (also known as Graves' ophthalmopathy) Atrial fibrillation Thyroid storm Pretibial myxedema (rare, non-pitting edema of the shin) Secondary hypogonadism (rare) Periodic thyrotoxic paralysis (rare) | Atrial fibrillation, congestive heart failure, thyroid eye disease, and thyroid storm are potential complications of Graves disease. |
| M36_Q3 | The presenting features of patients with hyperthyroidism may include:  Heat intolerance Sweating Tremor Palpitations Weight loss Fatigue Nervousness Irritability Rapid heartbeat Proximal muscle weakness Eyebrow loss Fine hair growth Thin skin Menstrual irregularities Diarrhea Goiter (enlarged thyroid) Thyroid eye disease (Graves disease) Lid retraction Proptosis (bulging eyes) Diplopia (double vision) These symptoms are not specific to hyperthyroidism and can be seen in other conditions as well. An accurate diagnosis can only be made after a thorough evaluation by a healthcare provider. | The presenting features of patients with hyperthyroidism include:  Heat intolerance Weight loss Palpitations Tremor Sweating Anxiety/nervousness Irritability Diarrhea Menstrual irregularities Muscle weakness Insomnia Increased appetite Goiter Thyroid eye disease (exophthalmos) | The following are presenting features of patients with hyperthyroidism: -Heat intolerance -Sweating -Weight loss -Tremor -Palpitations -Anxiety -Irritability -Insomnia -Menstrual irregularities -Goiter -Heat sensitivity -Increased appetite -Fatigue -Hyperactivity -Eyelid retraction -Exophthalmos -Increased muscle weakness. |

Raw ChatGPT output for each question posed within each vignette. “Question ID” is comprised of the assigned vignette number as a prefix, and the question type plus its count within the vignette as a suffix. *Misc* question IDs have the suffix “Q.”
